# IMMUNO-COV™ v2.0: Development and Validation of a High-Throughput Clinical Assay for Measuring SARS-CoV-2-Neutralizing Antibody Titers

**DOI:** 10.1101/2021.02.16.21251653

**Authors:** Rianna Vandergaast, Timothy Carey, Samantha Reiter, Chase Lathrum, Patrycja Lech, Clement Gnanadurai, Michelle Haselton, Jason Buehler, Riya Narjari, Luke Schnebeck, Anne Roesler, Kara Sevola, Lukkana Suksanpaisan, Alice Bexon, Shruthi Naik, Bethany Brunton, Scott C. Weaver, Grace Rafael, Sheryl Tran, Alina Baum, Christos A. Kyratsous, Kah Whye Peng, Stephen J. Russell

## Abstract

Neutralizing antibodies are key determinants of protection from future infection, yet well-validated high-throughput assays for measuring titers of SARS-CoV-2-neutralizing antibodies are not generally available. Here we describe the development and validation of IMMUNO-COV™ v2.0 a scalable surrogate virus assay, which titrates antibodies that block infection of Vero-ACE2 cells by a luciferase-encoding vesicular stomatitis virus displaying SARS-CoV-2 spike glycoproteins (VSV-SARS2-Fluc). Antibody titers, calculated using a standard curve consisting of stepped concentrations of SARS-CoV-2 spike monoclonal antibody, correlated closely (p < 0.0001) with titers obtained from a gold-standard PRNT50% assay performed using a clinical isolate of SARS-CoV-2. IMMUNO-COV™ v2.0 was comprehensively validated using data acquired from 242 assay runs performed over seven days by five analysts, utilizing two separate virus lots, and 176 blood samples. Assay performance was acceptable for clinical use in human serum and plasma based on parameters including linearity, dynamic range, limit of blank and limit of detection, dilutional linearity and parallelism, precision, clinical agreement, matrix equivalence, clinical specificity and sensitivity, and robustness. Sufficient VSV-SARS2-Fluc virus reagent has been banked to test 5 million clinical samples. Notably, a significant drop in IMMUNO-COV™ v2.0 neutralizing antibody titers was observed over a six-month period in people recovered from SARS-CoV-2 infection. Together, our results demonstrate the feasibility and utility of IMMUNO-COV™ v2.0 for measuring SARS-CoV-2-neutralizing antibodies in vaccinated individuals and those recovering from natural infections. Such monitoring can be used to better understand what levels of neutralizing antibodies are required for protection from SARS-CoV-2, and what booster dosing schedules are needed to sustain vaccine-induced immunity.

## INTRODUCTION

On March 11, 2020, the World Health Organization declared COVID-19, caused by SARS-CoV-2, a pandemic. Since then, the coordinated efforts of numerous researchers, biotechnology and pharmaceutical companies, contract manufacturers, healthcare organizations, and governmental agencies have resulted in the approval and initial distribution of the first SARS-CoV-2 vaccines. Clinical trial data indicate that the vaccines currently approved in the US are approximately 95% effective at preventing COVID-19 (1, 2). However, the durability of this protection is unknown. Neutralizing antibody responses following vaccination correlate with protective immunity (3–6), yet an increasing number of studies, including this one, demonstrate that neutralizing antibody levels fall steadily in the months following natural SARS-CoV-2 infection or vaccination (7–11). Thus, protective antibody responses, including those elicited by vaccination, may be relatively short-lived, and repeat vaccine dosing over several years may be necessary to achieve and maintain herd immunity. It is not currently known what titer of neutralizing antibodies confers protection from SARS-CoV-2 infection or COVID-19. Studies to monitor neutralizing antibody responses and the associated risk of infection at various timepoints post-vaccination are needed to inform decisions on the appropriate timing of booster vaccine doses. To facilitate these studies, a reliable, high-throughput method for quantitatively measuring neutralizing antibody titers is critically needed.

Over the course of the past year, numerous rapid serological tests have been developed, and many have received Emergency Use Authorization (EUA) approvals for the detection of antibodies against SARS-CoV-2. These tests, which are primarily enzyme-linked immunosorbent assay (ELISA)-based, provide a convenient way to identify individuals previously infected with SARS-CoV-2. However, it is well-known that only a small subset of virus-specific antibodies are capable of neutralizing virus infectivity, and thereby protecting against future viral infection and disease (12). Importantly, the rapid serological assays for which EUA approvals have been granted are not able to discriminate between neutralizing and non-neutralizing antibodies. Available evidence also suggests that post-vaccination and post-infection neutralizing antibody titers do not correlate strongly with total antibody titers (10,13–16), and it is unknown whether neutralizing antibody titers decay over time more rapidly than non-neutralizing antibodies. Thus, for reliable assessment of the level of protection against SARS-CoV-2 infection in vaccinated or previously infected individuals, neutralizing antibody assays are preferred.

The gold standard assay for the quantitation of virus neutralizing antibodies is the plaque-reduction neutralization test (PRNT). While providing a reasonable measure of the blood concentration of antibodies capable of neutralizing the SARS-CoV-2 virus, PRNT is labor intensive and requires use of a clinical virus isolate, such that the test can only be performed under biosafety level 3 containment. Safer alternative neutralization assays have been developed using non-replicating lentiviral vectors (10,14,17,18) or vesicular stomatitis viruses (VSVs)(19) pseudotyped with the SARS-CoV-2 spike glycoprotein. However, due to technical factors impacting the manufacture of these pseudotyped viruses, they are generally produced in small batches of variable titer, which significantly limits the scalability of these assays. The use of fully replication competent VSVs expressing the SARS-CoV-2 spike protein provides an attractive alternative for the development of neutralizing assays (20–22), as they can be propagated extensively to generate much larger reagent stocks. Moreover, because the natural VSV glycoprotein (G) is replaced with the SARS-CoV-2 spike protein, these recombinant viruses mimic SARS-CoV-2 entry, which is initiated by binding of the spike protein to its receptor angiotensin-converting enzyme 2 (ACE2) on the cell surface (23–25). Once bound to ACE2 via its receptor binding domain (RBD), the spike protein is proteolytically cleaved by the cell surface transmembrane serine protease TMPRSS2 or by endosomal cysteine proteases cathepsin B/L, providing a critical trigger for subsequent membrane fusion and virus entry into the cell (23, 26). Studies have mapped the targets of SARS-CoV-2-neutralizing antibodies to diverse epitopes within the spike protein, and antibodies that block ACE2 receptor binding, spike protein cleavage, or subsequent conformational rearrangements of the spike protein that lead to membrane fusion are all strongly neutralizing (27–31).

Here, we describe the development, optimization, and validation of IMMUNO-COV™ v2.0, a fully scalable neutralization assay that uses a replication competent G cistron-deleted recombinant VSV encoding both the SARS-CoV-2 spike protein and firefly luciferase (Fluc) (Fig. 1). Over 23,000 vials of this virus were prepared and cryopreserved from a single large-production run, providing sufficient material to assay more than 5 million serum or plasma samples. Anti-SARS-CoV-2-neutralizing antibody titers determined using IMMUNO-COV™ v2.0 demonstrated strong linear correlation with titers obtained using the classical PRNT under BSL-3. IMMUNO-COV™ v2.0 assay performance has remained robust and accurate for at least three months, during which time we have conducted extensive validation testing and subsequent verification studies. In keeping with the observations of other groups (7,8,16,28,32), higher titers of neutralizing antibodies were observed in subjects recovering from more severe SARS-CoV-2 infections, though strong responses were also seen in several subjects who had only mild disease symptoms. Importantly, a substantial decline in neutralizing antibody levels was observed in most COVID-19 convalescent subjects who were tested repeatedly over a six-month period, regardless of the initial antibody titer. Taken together, our results underscore the importance of monitoring neutralizing antibody titers of over time, and demonstrate how IMMUNO-COV™ v2.0 can be used to accurately quantify these responses at scale.

**Figure 1:**
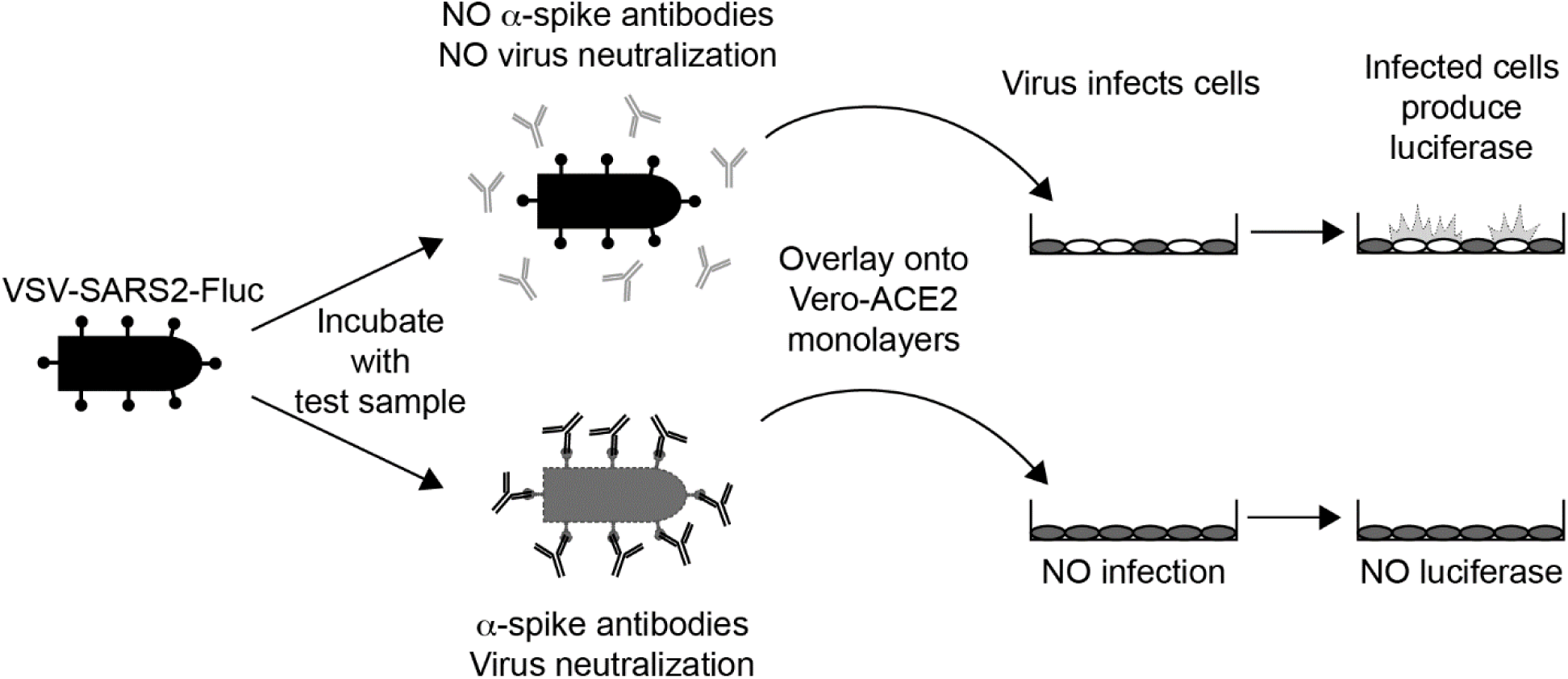
*Overview of the IMMUNO-COV™ v2.0 Assay.* A VSV expressing SARS-CoV-2 spike and firefly luciferase (VSV-SARS2-Fluc) is incubated with test sera or plasma. In the absence of SARS-CoV-2-neutralizing antibodies (top) the virus retains infectivity and infects Vero-ACE2 monolayers. If the test sample contains SARS-CoV-2-neutralizing antibodies (bottom), the antibodies inhibit infection by blocking cell entry. As virus replication proceeds, infected cells express luciferase, which is used to quantitate virus-infection. High luciferase signal means the test sample did not neutralize the virus, while decreased luciferase indicates the presence of SARS-CoV-2-neutralizing antibodies.

## Results

### Generation of VSV-SARS2-Fluc

Our previously published SARS-CoV-2 neutralization assay relied upon virus-induced fusion of two dual split protein (DSP) reporter cell lines to generate a luciferase signal (21). To further improve assay throughput and eliminate the need for two cell lines we generated a recombinant VSV (VSV-SARS2-Fluc) encoding SARS-CoV-2 spike-Δ19CT (S-Δ19CT) in place of VSV-G, and firefly luciferase (Fluc) as an additional transcriptional unit located between the S-Δ19CT and VSV-L genes (Fig. 2A). Cells infected with VSV-SARS2-Fluc express the virus-encoded luciferase, which is used to measure the level of virus infection. Incorporation of SARS-CoV-2 spike protein into VSV-SARS2-Fluc virions was confirmed by immunoblot (Fig. 2B). VSV-SARS2-Fluc infection and replication were also dependent on cellular ACE2 expression. Robust VSV-SARS2-Fluc replication and virus-induced cell death were observed in Vero-ACE2 cells, which overexpress the SARS-CoV-2 receptor ACE2 (Fig. 2C and D), but not in hamster BHK-21 cells (Fig. 2C and E), which do not express human ACE2. The control virus VSV-Fluc, which encodes VSV-G, but not S-Δ19CT, efficiently infected and replicated in Vero-ACE2 and BHK-21 cells. Cellular luciferase activity specifically correlated to replication of the Fluc-expressing viruses (Fig. 2F and G), with loss of luciferase signal at later timepoints coinciding with the death of infected cultures. Together, these data confirmed functional VSV-G replacement with S-Δ19CT and efficient Fluc expression from the VSV-SARS2-Fluc virus.

**Figure 2:**
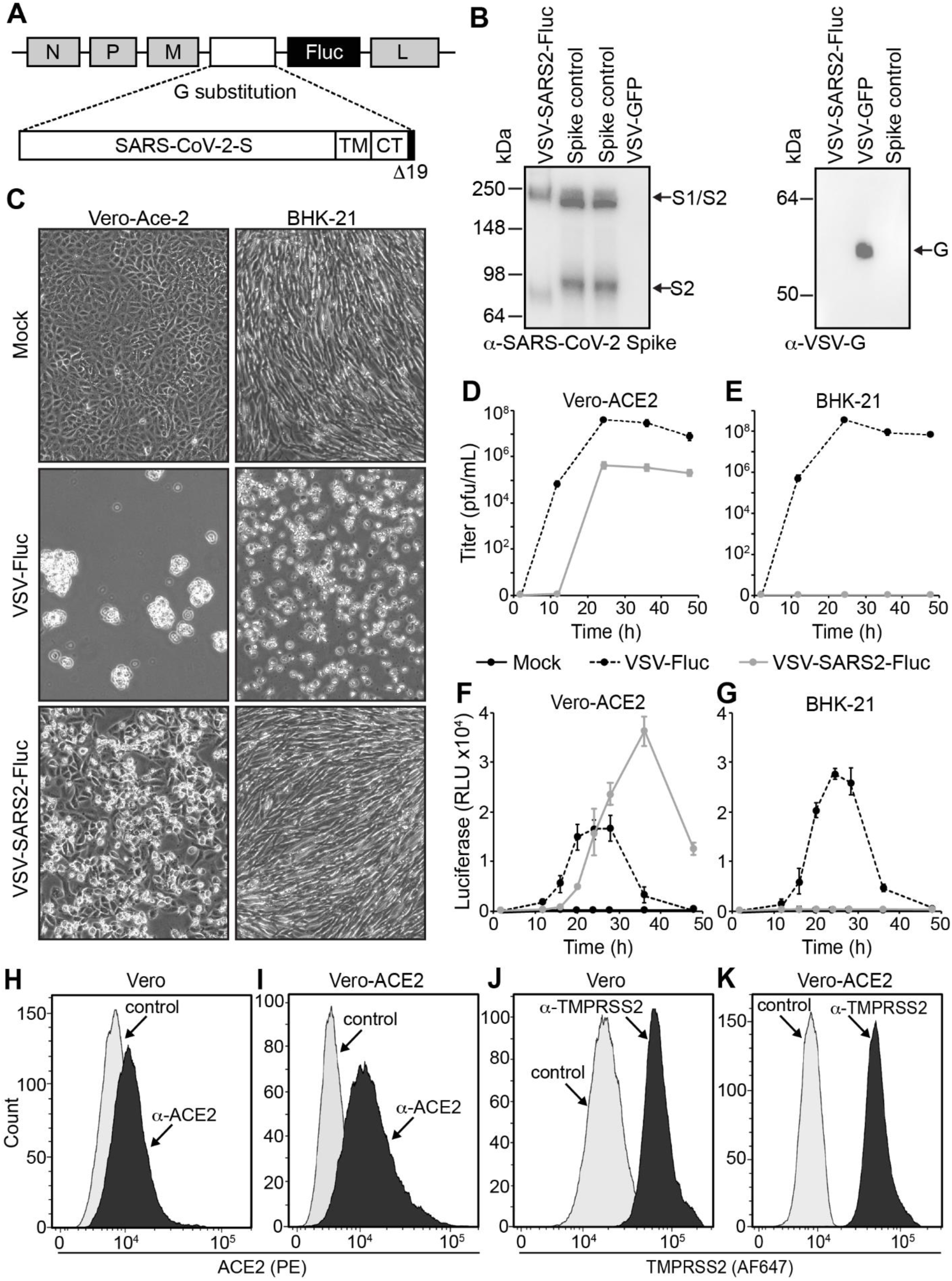
*Generation and Characterization of VSV-SARS2-Fluc.* A) Schematic Representation of the VSV-SARS2-Fluc Genome. The location of the VSV N, P, M (M51R), and L genes are shown. In place of VSV-G a codon optimized SARS-CoV-2 spike gene with a 19 amino acid C-terminal (CT) deletion (Δ19CT) is substituted. TM is transmembrane domain. Firefly luciferase (Fluc) is inserted as an additional transcriptional element between S-Δ19CT and L. Not drawn to scale. B) Immunoblot Analysis. VSV-SARS2-Fluc or VSV-GFP control virus (5 × 10^5^ total pfu) or spike control from lysates of cells overexpressing SARS-CoV-2 spike were subjected to immunoblot analysis using α-SARS-CoV-2 spike antibody (left) and α-VSV-G antiserum (right). Arrows indicate the full-length S1/S2 variant and cleaved S2 variant of spike and the VSV-G proteins. C) Infection of Cell Monolayers. Vero-ACE2 or BHK-21 cell monolayers were infected with VSV-SARS2-Fluc, control VSV-Fluc, or mock-infected. Images were taken 48 h post infection at 100X magnification. D-E) Replication Curves. Vero-ACE2 or BHK-21 cell monolayers were infected as in panel C and the virus titers from culture supernatants collected at the indicated times post inoculation were determined. F-G) Luciferase Activity. Vero-ACE2 or BHK-21 cells were infected with VSV-SARS2-Fluc, control VSV-Fluc, or mock-infected in 96-well plates, and at the indicated times luciferase activity was measured. H-K) Flow Cytometry. Expression of ACE2 (H and I) and TMPRSS2 (J and K) were measured in Vero and Vero-ACE2 cells by flow cytometry using α-ACE2 or α-TMPRSS2, respectively. Controls were secondary antibody only.

### Vero-ACE2 cells are an optimal cell substrate for detecting virus neutralization

VSV-SARS2-Fluc infects Vero cells via endogenously expressed ACE2 receptors (21). We hypothesized that ACE2 overexpression could enhance Vero cell susceptibility to VSV-SARS2-Fluc and thereby improve assay sensitivity. To this end, we tested Vero-ACE2 cells, which stably overexpress human ACE2 as confirmed by flow cytometry (Fig. 2H and I), in the assay. While Vero and Vero-ACE2 cells naturally express relatively high levels of TMPRSS2 (Fig. 2J and K), we also generated a stable cell line overexpressing both ACE2 and TMPRSS2 to elucidate the effect of TMPRSS2 on assay performance. VSV-SARS2-Fluc infection induced higher luciferase expression in Vero-ACE2 cells compared to Vero cells (Fig. 3A). Luciferase expression was not further enhanced by overexpression of TMPRSS2, and notably, VSV-SARS2-Fluc neutralization by the well-characterized neutralizing anti-SARS-CoV-2 spike monoclonal antibody mAb10914 was less apparent on Vero-ACE2/TMPRSS2 cells compared to Vero-ACE2 cells (Fig. 3A). Since Vero-ACE2 cells provided for more sensitive detection of viral neutralization, these cells were selected as the cell substrate for assay development.

**Figure 3:**
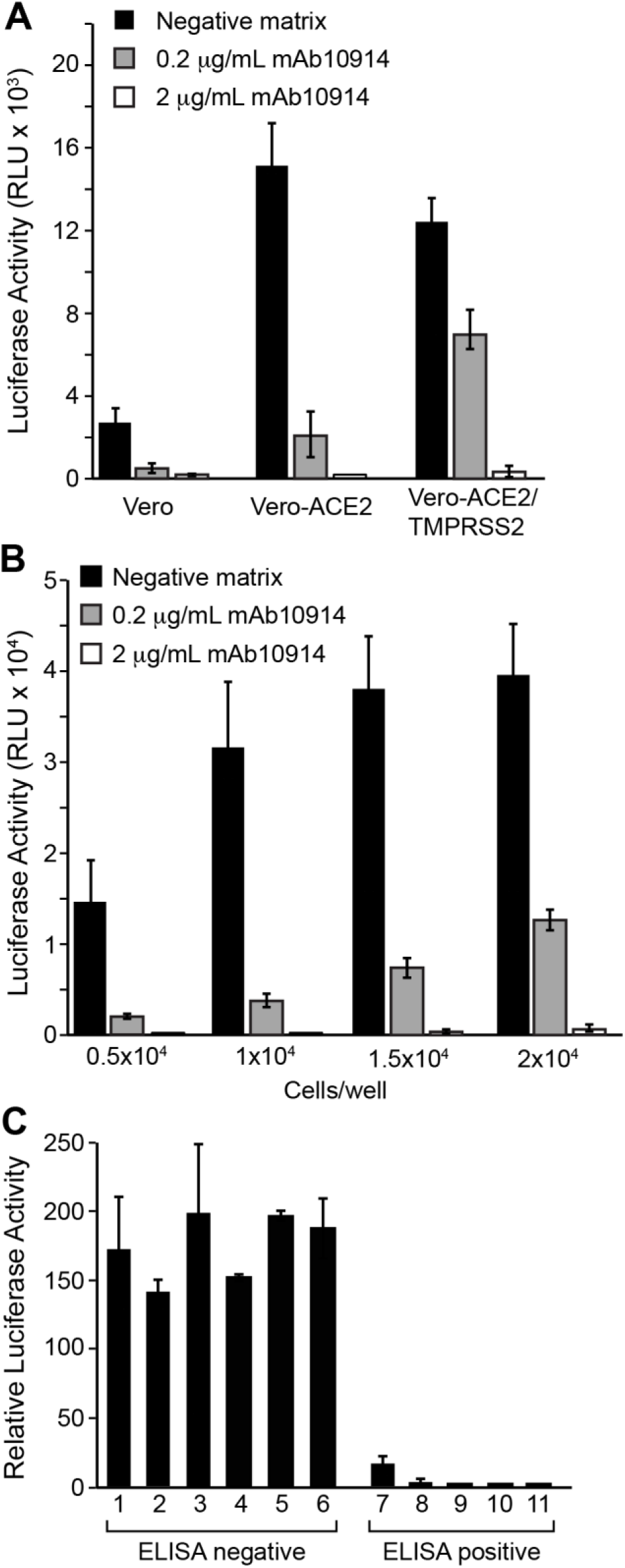
*Inhibition of VSV-SARS2-Fluc by Monoclonal Antibodies and Convalescent Sera.* A) Infectivity of Different Vero Cell Lines. VSV-SARS2-Fluc was incubated with 2 or 0.2 µg/mL of monoclonal anti-SARS-CoV-2 spike antibody mAb10914 in pooled seronegative sera, or pooled seronegative sera alone (negative matrix). After 30 min, virus mixes were overlaid onto Vero, Vero-ACE2, or Vero-ACE2/TMPRSS2 cells. Luciferase activity was measured after an additional 24 h. Values represent the average (mean) RLU ± standard deviation. B) Optimization of Cell Density. The indicated numbers of Vero-ACE2 cells were seeded in 96-well plates. The following day, virus mixes as described in panel A were overlaid onto the cell monolayers. Luciferase activity was measured after an additional 24 h. Values represent the average (mean) RLU ± standard deviation. C) Neutralization by Convalescent Sera. VSV-SARS2-Fluc was incubated with pooled seronegative sera at 1:80 dilution or sera samples from 11 donors (6 seronegative, 5 seropositive for anti-SARS-CoV-2 antibodies by ELISA assay) at 1:80 dilution. After 30 min, virus/sera mixes were overlaid onto Vero-ACE2 cells. Luciferase activity was measured after an additional 24 h. Values represent average (mean) luciferase activity relative to the pooled seronegative sera sample control ± standard deviation.

We also examined the effect of Vero-ACE2 cell seeding density on assay performance. Higher luciferase activity was detected when cell density was increased from 5,000 to 10,000 cells/well (96 well plate; Fig. 3B), but further increasing the cell density to 20,000 cells/well led to only a modest additional incremental increase in luciferase signal. Moreover, the higher cell density of 20,000 cells/well was associated with a less effective neutralization of luciferase signal when the virus was exposed to the neutralizing antibody mAb10914. We concluded that 10,000 cells/well was the optimal seeding density for detection of virus neutralization.

To demonstrate the detection of neutralizing antibodies in patient samples, we used serum samples confirmed as seronegative or seropositive by the commercial EUROIMMUN Anti-SARS-CoV-2 ELISA (IgG), which detects anti-SARS-CoV-2 spike antibodies. Serum samples were incubated with VSV-SARS2-Fluc for 30 minutes at room temperature then added to culture wells containing pre-plated Vero-ACE2 cells. All five of the seropositive samples substantially inhibited virus infection, resulting in suppression of luciferase activity (Fig. 3C). No reduction in luciferase activity was observed when the VSV-SARS2-Fluc virus was pre-incubated with seronegative samples, confirming that neutralizing antibodies were detected only in seropositive donor samples.

### Consistency of different VSV-SARS2-Fluc production lots

To determine the optimal quantity of virus to add to each assay well, we tested the capacity of mAb10914 and seropositive plasma to neutralize increasing amounts of VSV-SARS2-Fluc. Highly neutralizing seropositive plasma and mAb10914 at a concentration of 2 µg/mL inhibited infectivity by at least 90%, independent of the amount of virus added to the well (Fig. 4A). In contrast, mAb10914 at a concentration of 0.2 µg/mL noticeably blocked infectivity in this assay only when less than 900 plaque forming units (pfu) of virus were added to each well. Based on this experiment, the optimal quantity of VSV-SARS2-Fluc virus to be added to each well to ensure sensitive detection of low-levels of neutralizing antibodies is between 200 and 400 pfu. Consistency of virus lots was confirmed by comparing mAb10914 inhibition of two independent lots of VSV-SARS2-Fluc (produced at different times and representing subsequent virus passages). Luciferase activity over a range of concentrations of mAb10914 was nearly indistinguishable between the two different virus lots (Fig. 4B). Comparing the mAb10914 inhibition curve with 200, 300, and 400 pfu of virus per well, the linear range was slightly wider when 300 pfu/well of VSV-SARS2-Fluc was used. Therefore, we used 300 pfu for all future assay runs. We also tested the stability of the thawed VSV-SARS2-Fluc virus when stored on ice or at room temperature prior to being used in the assay. No significant reduction in virus infectivity or neutralization occurred following an 8-hour incubation on ice (Fig. 4C). Likewise, the virus was stable for up to an hour at room-temperature, with only a modest titer decrease noted after two hours (Fig. 4D).

**Figure 4:**
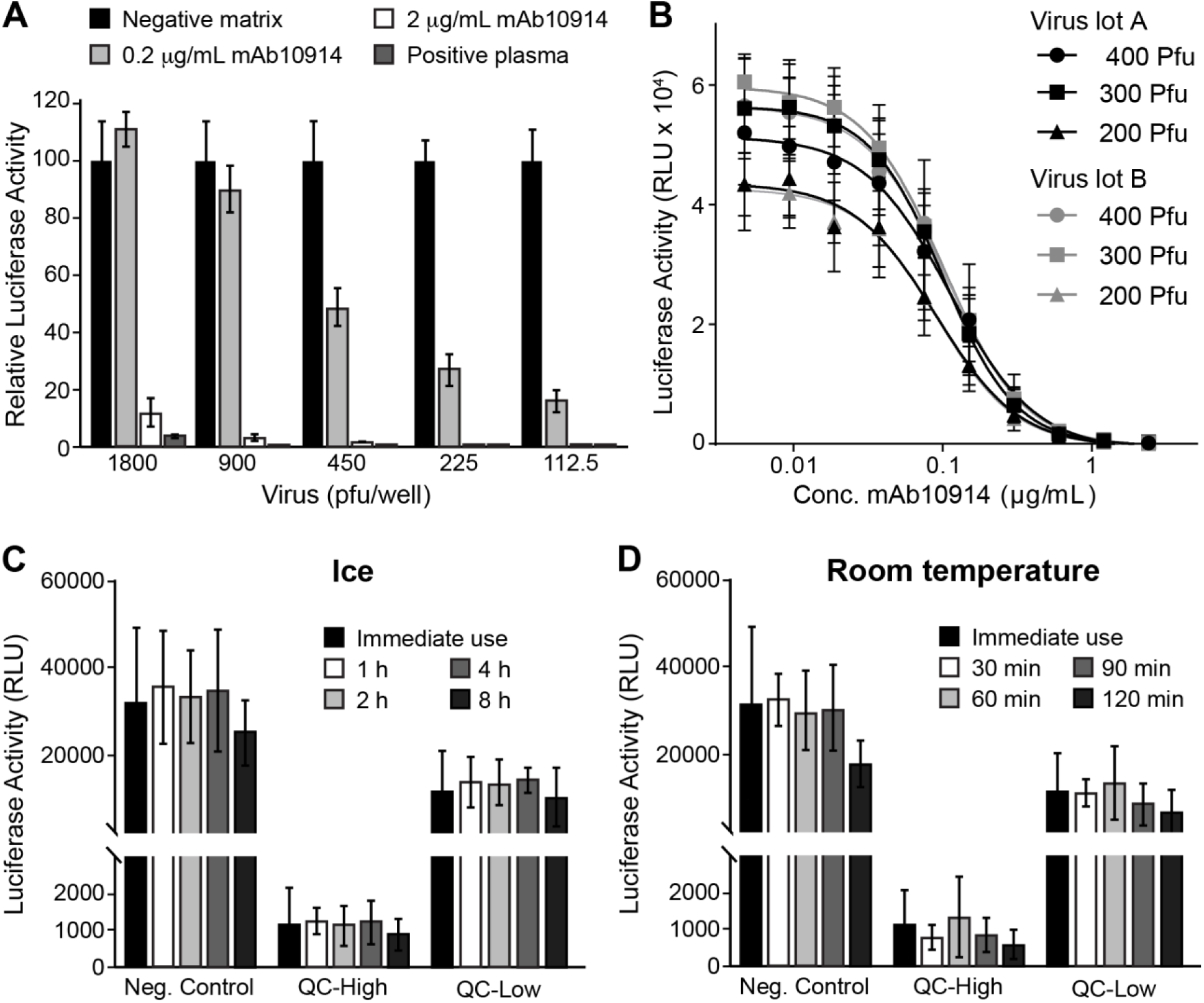
*Assay Performance of VSV-SARS2-Fluc.* A) Susceptibility of Virus to Antibody Neutralization. The indicated amounts (plaque forming units; pfu) of VSV-SARS2-Fluc were incubated with 2 or 0.2 µg/mL anti-SARS-CoV-2 spike monoclonal antibody mAb10914, a SARS-CoV-2 seropositive plasma sample at 1:80 dilution, or pooled seronegative serum (negative matrix, 1:80). After 30 min, virus mixes were overlaid onto Vero-ACE2 cells, and luciferase activity was measured after an additional 24 h. Values represent the average (mean) luciferase activity relative to the negative matrix control ± standard deviation. B) Consistency of Virus Lots. Varying amounts (pfu) of two different lots (A and B) of VSV-SARS2-Fluc were incubated with the indicated concentrations of mAb10914. Luciferase activity was measured after an additional 24 h. Values represent the average (mean) RLU ± standard deviation. C-D) Virus Stability. Aliquots of VSV-SARS2-Fluc were removed from the freezer, thawed, and either used immediately for assay (Immediate use) or stored at either room temperature or on ice for the indicated time (h). For assay, 300 pfu of VSV-SARS2-Fluc was incubated with 0.154 (QC-High) or 0.031 (QC-Low) µg/mL of anti-SARS-CoV-2 spike monoclonal antibody mAb10922 in pooled seronegative sera, or in pooled seronegative sera alone (Neg. Control). After 30 min, virus mixes were overlaid onto Vero-ACE2 cells and luciferase activity was measured after an additional 24 hours. Values represent the average (mean) RLU ± standard deviation.

### Heat-inactivation of serum samples is not necessary for assay compatibility

In cellular assays, heat-inactivation of plasma and serum samples is often necessary to limit matrix interference that can affect cell or virus viability. To determine whether heat-inactivation was required for IMMUNO-COV™ v2.0, twenty matched serum and plasma samples were thawed and aliquoted, with one aliquot kept on ice, while the other aliquot was heat-inactivated at 56°C for 30 minutes. Both aliquots were then tested in the assay. Overall, heat-inactivation had little effect on neutralizing activity. All seronegative samples remained negative and all seropositive samples remained positive in the assay, regardless of whether the samples had been heat-inactivated (Fig. 5A and 5B). Importantly, heat-inactivated samples did not exhibit diminished virus neutralizing capacity, suggesting that complement proteins do not enhance the neutralization of VSV-SARS2-Fluc in this assay format. For plasma samples, heat-inactivation and subsequent clarification prevented thermal coagulation and sample loss during the assay, thereby improving assay performance. We therefore continued to use heat-inactivation for all subsequent assays with plasma samples, while using non-heat-inactivated serum samples.

**Figure 5:**
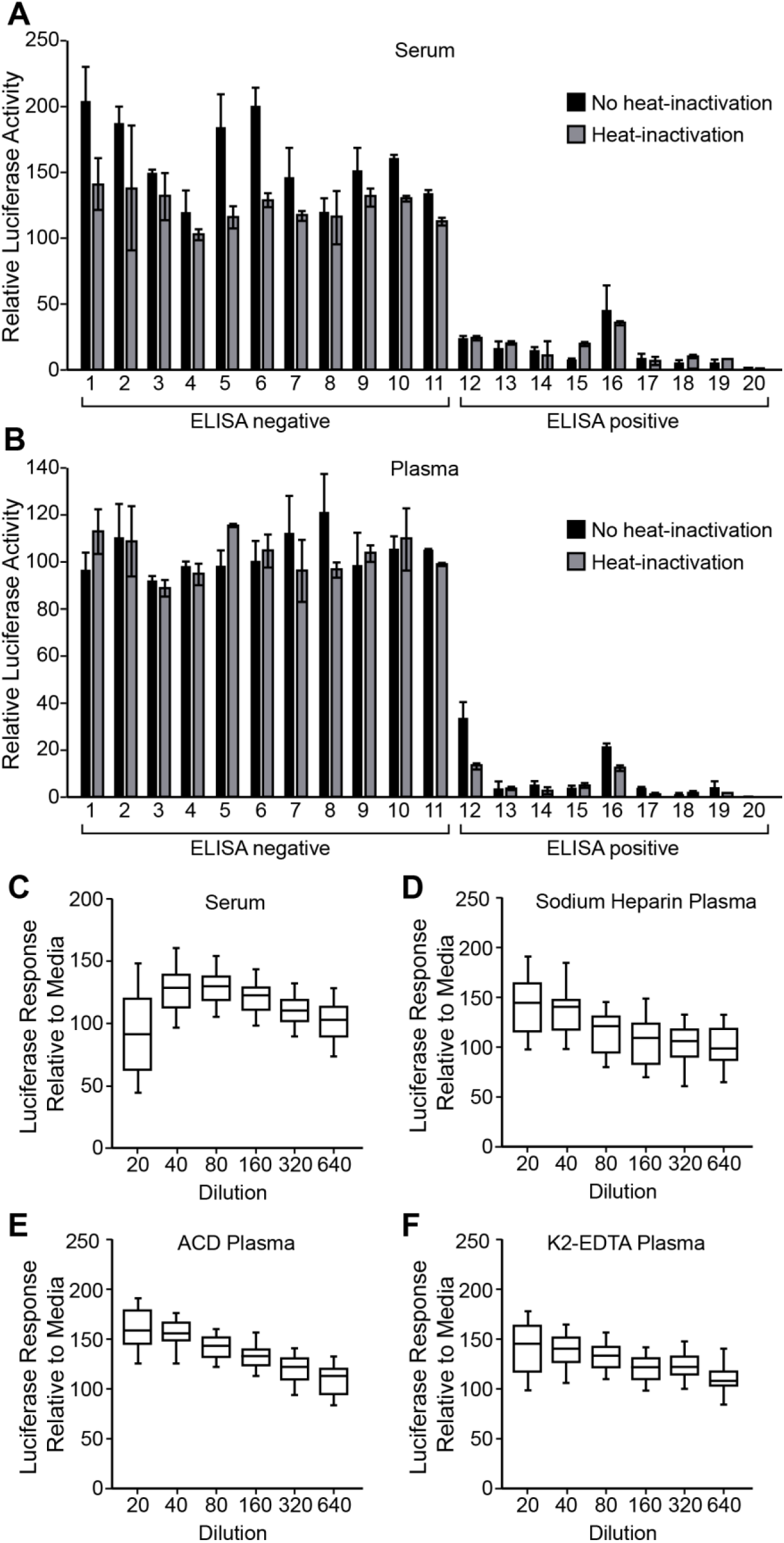
*Effect of Sample Matrix on Assay Performance*. A-B) Effect of Heat-Inactivation of Sera or Plasma. Matched sera (A) and sodium-heparin plasma (B) samples from 20 donors (11 seronegative, 9 seropositive for anti-SARS-CoV-2 antibodies by ELISA assay) were split and either incubated on ice or at 56°C for 30 min. Following incubation, plasma samples were clarified by centrifugation. Samples were then incubated at 1:80 dilution with VSV-SARS2-Fluc. Pooled seronegative sera or plasma were used as assay controls. After 30 min, virus mixes were overlaid onto Vero-ACE2 cells, and after an additional 24 h, luciferase activity was measured. Values represent the average (mean) luciferase activity relative to the pooled seronegative matrix control ± standard deviation. C-F) Characterization of Matrix Interference. Seronegative sera (C, n=40), sodium-heparin plasma (D, n=40), ACD plasma (E, n=26), or K2-EDTA plasma (F, n=49) samples were serially diluted as indicated and incubated with VSV-SARS2-Fluc. Virus mixed with media only was used as a control. After 30 min, virus mixes were overlaid onto Vero-ACE2 cells, and after an additional 24 h, luciferase activity was measured. Values represent the average (mean) luciferase activity relative to the media control ± standard deviation.

### Serum and plasma demonstrate low matrix interference

In our original cell fusion-based IMMUNO-COV™ assay we observed significant matrix interference at high concentrations of serum and plasma (21). To determine whether IMMUNO-COV™ v2.0, which provides a more direct measure of virus infection, is similarly hampered by matrix interference, we ran numerous seronegative samples in the assay at 2-fold serial dilutions ranging from 1:20 through 1:640. Minimal matrix interference was observed with serum, sodium-heparin plasma, ACD plasma, and K2-EDTA plasma (Fig. 5C, D, E, and F). In fact, higher concentrations of plasma appeared to have a stabilizing effect on the virus relative to cell culture medium alone and were associated with higher levels of luciferase activity at assay readout. Likewise, serum appeared to increase virus stability relative to medium alone, though some matrix interference was observed at the 1:20 dilution. Thus, the IMMUNO-COV™ v2.0 assay is compatible with testing at low sample dilutions, which may be of importance if higher detection sensitivities are desired.

### Quantification of neutralizing antibody titers using a standard curve

To determine the titer of neutralizing antibodies in a test sample without the need for serial two-fold sample dilutions, we developed an assay format in which just one or two dilutions of a test sample are read against a standard calibration curve included on every assay plate. For the development of a calibration standard and assay controls, we used two well-characterized neutralizing anti-spike monoclonal antibodies, mAb10914 and mAb10922. Both antibodies neutralized VSV-SARS2-Fluc in a dose-dependent manner (Fig. 6A), whereas no virus inhibition was observed using isotype antibody at any of the concentrations tested. Based on these findings we established a six-point standard curve using two-fold dilutions of mAb10914 in tissue culture medium at concentrations ranging from 0.8 µg/mL to 0.025 µg/mL (Fig. 6B). To quantify the viral neutralizing titers of test samples, each antibody concentration in the standard curve was converted to a virus neutralizing titer (VNT) by multiplying the antibody concentration by 400. The correction factor of 400 was chosen as it produced VNT values that approximated PRNT50% values obtained for samples assayed at a 1:80 dilution (see below). The final standard curve range for the assay therefore gives a VNT readout of 10-320 for a sample assayed at a 1:80 dilution. In numerous tests (n=242 assay runs), the 160, 80, 40, and 20 VNT standards fell within the linear range >99% of the time (Table 1). In most runs (87.6%), either the 320 or 10 VNT standard was also within the linear range. Thus, the standard curve effectively spanned the assay linear range. To quantitate antibody titers above 320 VNT, additional sample dilutions above 1:80 were employed in the assays described below.

**Figure 6:**
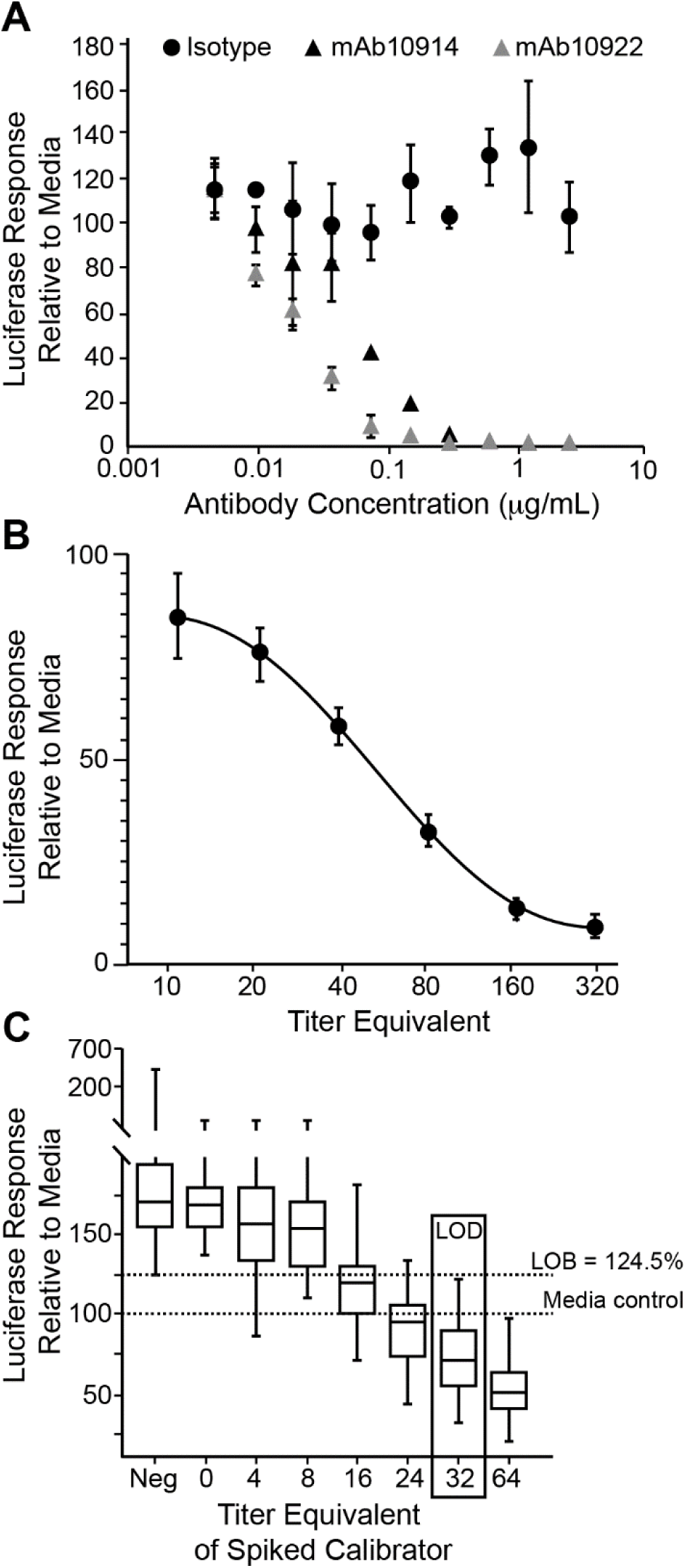
*Establishment of a Standard Curve for Titer Calculations.* A) Antibody-Specific Neutralization of VSV-SARS2-Fluc. The indicated concentrations of anti-SARS-CoV-2 spike monoclonal antibodies mAb10914 or mAb10922 or isotype control antibody were incubated with VSV-SARS2-Fluc. After 30 min, virus mixes were overlaid onto Vero-ACE2 cells, and luciferase activity was measured after an additional 24 h. Values represent the average (mean) luciferase activity relative to the media control ± standard deviation. B) Standard Curve Performance. VSV-SARS2-Fluc was incubated with 0.8, 0.4, 0.2, 0.1, 0.05, or 0.025 µg/mL (corresponding to the indicated equivalent VNTs) of mAb10914 or negative pooled sera alone. After 30 min, virus mixes were overlaid onto Vero-ACE2 cells, and luciferase activity was measured after an additional 24 h. Values represent average (mean) luciferase activity relative to the pooled negative sera control ± standard deviation from 242 unique assay runs. C) Limit of Detection. Five different seronegative sera samples (at 1:80 dilution) were spiked with anti-SARS-CoV-2 spike monoclonal antibody mAb10914 at 0.01, 0.02, 0.04, 0.06, 0.08, and 0.1 µg/mL (corresponding to the indicated equivalent VNTs), and incubated with VSV-SARS2-Fluc. VSV-SARS2-Fluc incubated with unspiked sera samples (Neg) or media alone were included as controls. After the 30 min incubation, virus mixes were overlaid onto Vero-ACE2 cells, and luciferase activity was measured after an additional 24 h. Box and whisker diagrams display the interquartile range in the box, with the center line representing the median for the data set and whiskers representing the lower 5% and upper 95% value. Values are based on a total of 12 different assay runs performed on three separate days by six analysts using two different virus lots.

**Table 1:**
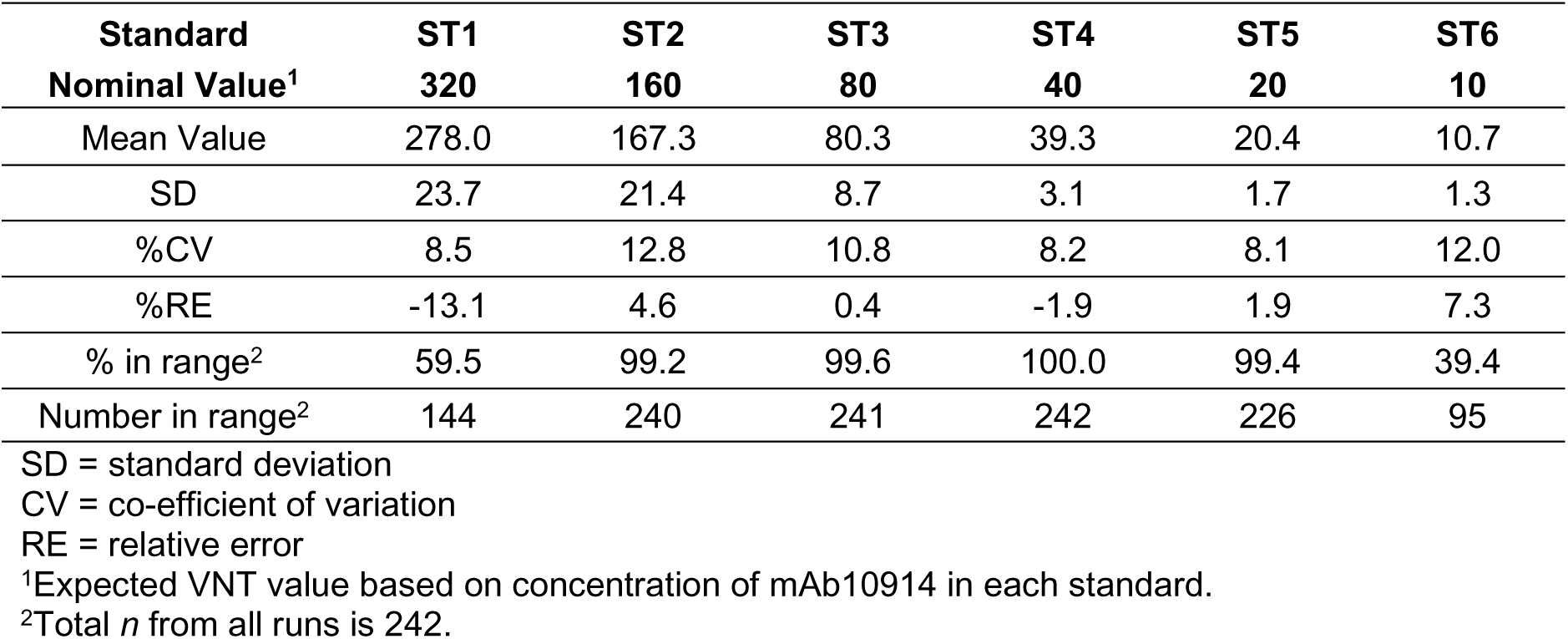
Assay Linearity

### Under standard conditions the assay limit of detection is 32 VNT

To determine the limit of detection (LOD) of the assay we first determined the assay limit of blank (LOB), representing the background signal from seronegative serum. To this end, we assayed seven known seronegative serum samples at a 1:80 dilution on 12 assay runs and calculated the luciferase signal as a percentage of the signal in media only controls. As observed previously (Fig. 5C), seronegative samples stabilized virus, and the LOB was a luciferase response of 124.5% compared to media alone. Seronegative serum samples were subsequently spiked with low concentrations of standard mAb10914 (0.01, 0.02, 0.04, 0.06, 0.08, and 0.1 µg/mL, corresponding to VNTs of 4, 8, 16, 24, 32, and 40) and assayed side-by-side with unspiked samples (Fig. 6C). Based on a total of 60 values obtained for each spike level, the lowest concentration of mAb10914 at which ≥95% of the luciferase response values were below the LOB was 0.08 µg/mL. This concentration corresponded to a VNT of 32, which was accepted as the LOD for the assay.

### The assay exhibits high specificity and sensitivity

To evaluate the sensitivity and specificity of IMMUNO-COV™ v2.0 when used to discriminate between positive and negative results, we performed blinded testing of 176 serum samples that were categorized as either positive or negative for SARS-CoV-2-neutralizing antibodies based on the readouts from ELISA and gold standard PRNT. All samples that tested positive for SARS-CoV-2 spike binding antibodies by ELISA were subsequently analyzed by PRNT, with only those samples that were positive by PRNT considered positive for neutralizing antibodies. Samples that tested negative by ELISA but positive in the IMMUNO-COV™ v2.0 assay were also tested by PRNT to confirm the presence or absence of neutralizing antibodies. In these analyses, our assay demonstrated 100% specificity when compared to both PRNT50% and PRNT80% results, as all PRNT-negative samples tested negative in IMMUNO-COV™ v2.0 (Table 2). Assay sensitivity was 93.7% relative to PRNT50% and 98.4% relative to PRNT80%. Moreover, 140 serum samples acquired prior to March 2020 (134 acquired from 2017-2019, 5 acquired in early 2020) from donors recovered from infection with endemic human coronaviruses HKU1 (n=35), NL63 (n=32), OC43 (n=35), or 229E (n=35) were all negative for neutralizing antibodies when tested using the IMMUNO-COV™ v2.0 assay. Thus, the assay specifically detected neutralizing antibodies to SARS-CoV-2 and most likely does not cross react to the four common human coronaviruses.

**Table 2:** Assay Specificity and Sensitivity

|  | Specificity<sup>1</sup> | Sensitivity<sup>2</sup> |
| --- | --- | --- |
| **PRNT50%** |  |  |
| Percentage | 100% | 93.7% |
| Sample Agreement | 113/113 | 59/63 |
| **PRNT80%** |  |  |
| Percentage | 100% | 98.4% |
| Sample Agreement | 116/116 | 59/60 |

<sup>1</sup>IMMUNO-COV™ v2.0 negative results relative to PRNT/ EUROIMMUN IgG ELISA negative results (samples positive by ELISA but negative by PRNT were considered negative).
<sup>2</sup>IMMUNO-COV™ v2.0 positive results relative to PRNT positive results. Any samples collected from donors previously PCR-positive for SARS-CoV-2, or positive for SARS-CoV-2 antibodies by IMMUNO-COV™ v2.0 or EUROIMMUN IgG ELISA were tested by PRNT assay.

We also assessed assay variability. Each of the blinded serum samples was assayed on five distinct runs performed by four different operators over a period of five days. Perfect consensus of positive and negative results between all five runs was observed for 174 (98.9%) of the samples. Antibody titers of positive samples were consistent between operators and assay runs, with titers across five different runs exhibiting 27.9% CV (n=59), which compared favorably to a CV of 65.1% for the PRNT (n=8 samples, two separate runs). Inter-assay precision was also evaluated based on the performance of the standard curve and assay controls. For this purpose, we included quality control (QC) high (0.154 µg/mL) and QC low (0.031 µg/mL) controls consisting of mAb10922 spiked into negative pooled sera on each assay plate. From 207 assay runs, QC high and QC low VNT readouts both demonstrated less than 30% inter-assay variability (Table 3 and Fig. 7). Intra-assay variability, which was assessed by running the same controls in 24 wells of the same plate, was below 20% for both controls (QC high = 8.6%, QC low = 19.1%). Collectively, these data demonstrate that the IMMUNO-COV™ v2.0 assay has acceptably low levels of intra- and inter-assay variability.

**Figure 7:**
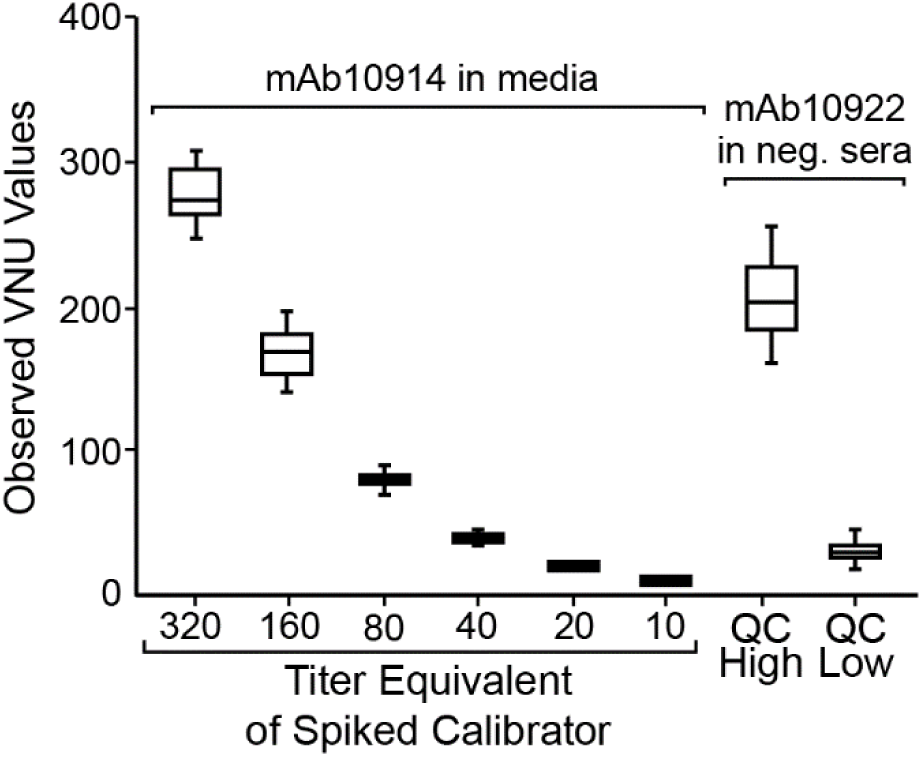
*Inter-Assay Variability of Standards and Controls.* Standards consisting of monoclonal anti-SARS-CoV-2 spike antibody mAb10914 at 0.8, 0.4, 0.2, 0.1, 0.05, and 0.025 µg/mL in media, and QC High and QC Low controls consisting of 0.154 and 0.031 µg/mL antibody mAb10922 in pooled seronegative sera were incubated with VSV-SARS2-Fluc. Pooled seronegative sera alone was used as a negative control. After 30 min, virus mixes were overlaid onto Vero-ACE2 cells, and luciferase activity was read after an additional 24 h. A total of 207 assay runs were performed over five days, by five analysts, using two different virus lots. Box plot represents the 25^th^ to 75^th^ percentile of the data with the line representing the media titer equivalent (VNT) value. Whiskers display the minimum and maximum values.

**Table 3:**
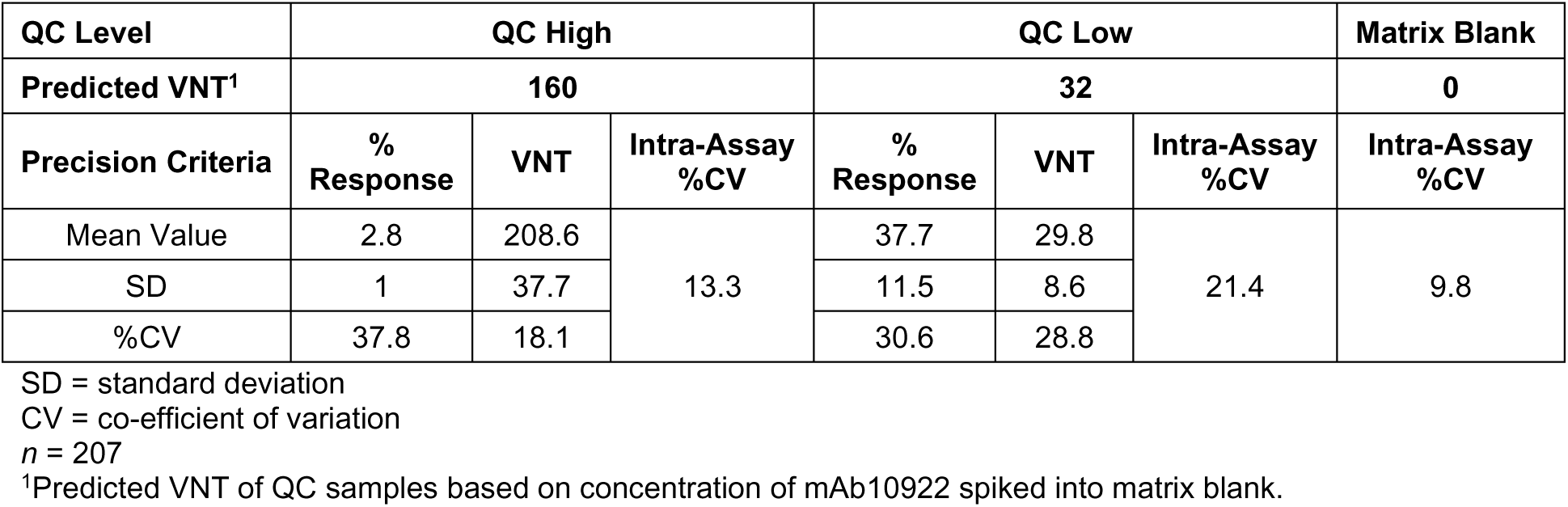
Intra- and Inter-Assay Variability

### Assay equivalence of serum and plasma samples

While most of our assay validation studies were conducted using serum samples, we also performed matrix equivalency testing to confirm assay compatibility with different plasma matrices. To this end, we acquired matched serum, sodium heparin plasma, ACD plasma, and K2/EDTA plasma samples from 26 of the 176 subjects whose serum samples were used to evaluate assay specificity and sensitivity, and tested the matched samples side-by-side in the assay. The consensus results and VNT antibody titers of positive samples from five assay runs were compared for each matrix. The average percentage relative error for each matrix was within ±30% for all plasma matrices (Table 4). Although all three plasma matrices demonstrated equivalency in this experiment, in other experiments (data not shown) the sodium heparin plasma samples did not exhibit dilutional linearity. Thus, only ACD plasma and K2/EDTA plasma are currently considered acceptable matrices for clinical testing.

**Table 4:** Matrix Equivalency

|  | Sodium<br>heparin-plasma | ACD-plasma | K2/EDTA-plasma |
| --- | --- | --- | --- |
| **%RE**<br>(relative to sera)<sup>1</sup> | +9.3 | -9.8 | +24.6 |

%RE = percent relative error
<sup>1</sup>Consensus VNT titers from the indicated plasma samples were compared to the consensus VNT titer for the matched serum sample. $n=12$ .

### IMMUNO-COV™ v2.0 VNT antibody titers correlate closely to PRNT50% titers

The BSL-3 PRNT with wild-type SARS-CoV-2 remains the gold standard for detection of neutralizing antibodies. Therefore, we compared the titers (VNT) measured using IMMUNO-COV™ v2.0 with those determined by PRNT. A strong correlation (Pearson’s R = 0.8870, p < 0.0001) was observed between VNTs and PRNT50% titers (Fig. 8). Therefore, neutralization of VSV-SARS2-Fluc in our assay closely mirrors the neutralization of SARS-CoV-2, and IMMUNO-COV™ v2.0 titers provide an accurate measure of an individual’s level of neutralizing antibodies. Moreover, VNTs can be quickly compared to PRNT50% titers using a conversion table (Table 5), which we generated based on our data obtained using the two different assays.

**Figure 8:**
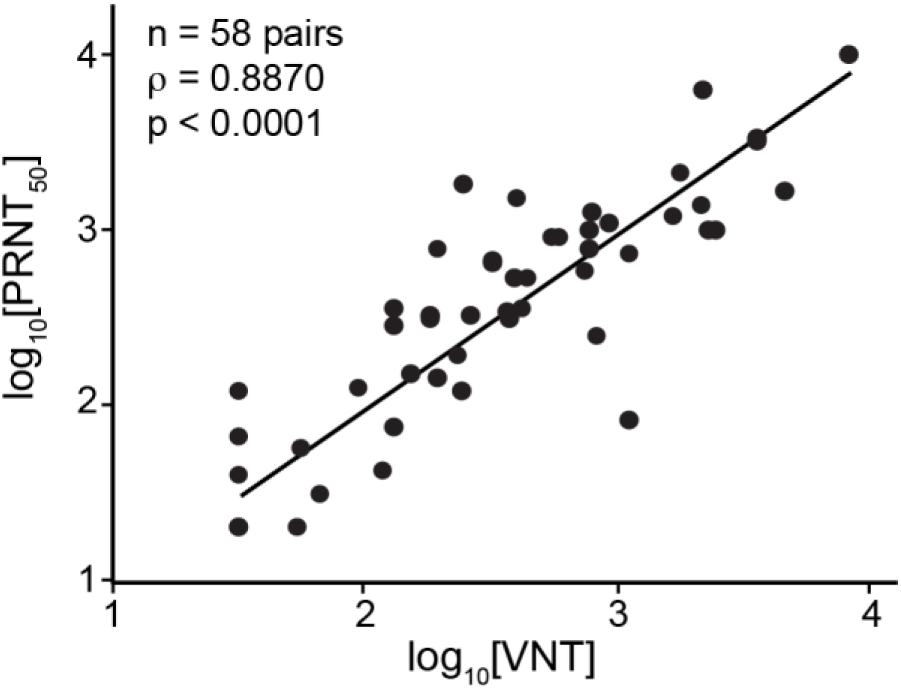
*Correlation of Virus Neutralizing Units to PRNT50%.* 58 SARS-CoV-2 seropositive sera samples were assayed using IMMUNO-COV™ v2.0 starting at a 1:80 dilution. Established controls, including a standard curve (0.8, 0.4, 0.2, 0.1, 0.05, and 0.025 µg/mL mAb10914 in media), were included on each assay plate. The IMMUNO-COV™ v2.0 titer (VNT) was determined using the standard curve, where one VNT equals the concentration of mAb10914 multiplied by 400. All samples were subjected to PRNT using a clinical isolate of SARS-CoV-2. Statistical comparison of VNT relative to PRNT50% was performed using Spearman’s rank order correlation analysis as both datasets had a non-gaussian distribution (*p* < 0.0001).

**Table 5:** VNT to PRNT50% Conversion

| VNT | PRNT50% |
| --- | --- |
| < 32 | < 1:40 |
| 32 to 40 | 1:40 |
| 41 to 80 | 1:80 |
| 81 to 180 | 1:160 |
| 181 to 400 | 1:320 |
| 401 to 800 | 1:640 |
| 801 to 1600 | 1:1280 |
| 1601 to 2400 | 1:2560 |
| > 2400 | > 1:2560 |

### Individuals with more severe disease symptoms tend to develop higher titers of neutralizing antibodies

Increasing evidence indicates that disease severity influences the strength of the neutralizing antibody response (7,8,16,28,32). To examine whether individuals in our study with more severe disease developed higher titers of neutralizing antibodies, we correlated antibody titers with self-reported disease symptoms from 46 previously infected donors who had tested positive for SARS-CoV-2-neutralizing antibodies. Samples used for this analysis were collected within the time window of two weeks to two months post confirmation of COVID-19 diagnosis. A wide range of neutralizing antibody titers was observed among these donors with significant overlap between the disease severity groupings (Fig. 9). Mean neutralizing antibody titers increased with increasing disease severity, though differences were not statistically significant. Our data, therefore, support previous findings that strong neutralizing antibody responses are more likely in individuals who have recovered from severe disease, but wide variation in neutralizing titers occur within all disease severity groupings.

**Figure 9:**
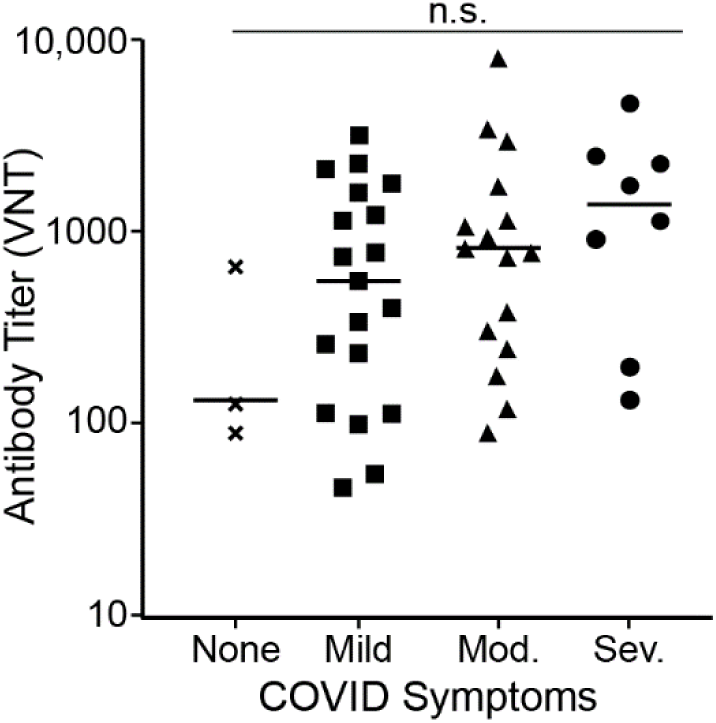
*The* S*trength of Neutralizing Antibody Responses Correlates to Disease Severity*. As part of assay validation (Table 2), neutralizing antibody titers were determined for 46 donors who self-reported COVID-19 disease symptoms at least two weeks prior to sample donation. Disease symptoms were classified as severe (acute respiratory distress or pneumonia), moderate (shortness of breath), mild (fever, feverish, cough, chills, myalgia, rhinorrhea, sore throat, nausea/vomiting, headache, abdominal pain, or diarrhea), or none (asymptomatic). The graph indicates the titer value (VNT) for each donor grouped based on disease symptoms. Bars represent the average (mean) titer for each group. Differences in antibody titers based on disease severity were not statistically significant (n.s.) by one-way ANOVA (*p* = 0.1904).

### SARS-CoV-2-neutralizing antibody titers fall steadily after recovery from infection

To provide long-term protection from COVID-19, neutralizing antibodies must persist at sufficiently high levels to block infection or mitigate pathogenesis. To examine the durability of SARS-CoV-2-neutralizing antibodies after recovery from natural infection, we determined the change in neutralizing antibody titers from 13 subjects between April and October 2020. In April, all 13 of these subjects had been diagnosed with COVID-19 within the previous two months and had measurable levels of SARS-CoV-2-neutralizing antibodies. Samples collected in April were stored at ≤ -65°C and assayed side-by-side with new samples collected in October from the same subjects. A two- to five-fold drop in neutralizing antibody titers was observed in all but one subject (Fig. 10A and Table 6). The outlier showed a 3-fold increase, suggesting possible asymptomatic re-exposure to the virus. In three subjects, the VNT from October dropped below the limit of detection in serum, though neutralizing antibodies could still be detected at very low levels in ACD-plasma from two of these subjects. Together, these data indicate that SARS-CoV-2 neutralizing antibody titers fall quite rapidly over time following natural infection. Importantly, while the PRNT confirmed the substantial decrement in SARS-CoV-2-neutralizing antibody titers over six months (Fig. 10B), a similar trend was not observed using a “neutralization” assay that measures binding of the spike RBD to immobilized ACE2 receptor (Fig. 10C and Table 6). When samples were tested using this SARS-CoV-2-spike RBD binding assay, antibody levels in several subjects were similar in April and October. This finding highlights the importance of quantifying neutralizing antibodies by inhibition of live virus rather than relying on a surrogate receptor binding assay.

**Figure 10:**
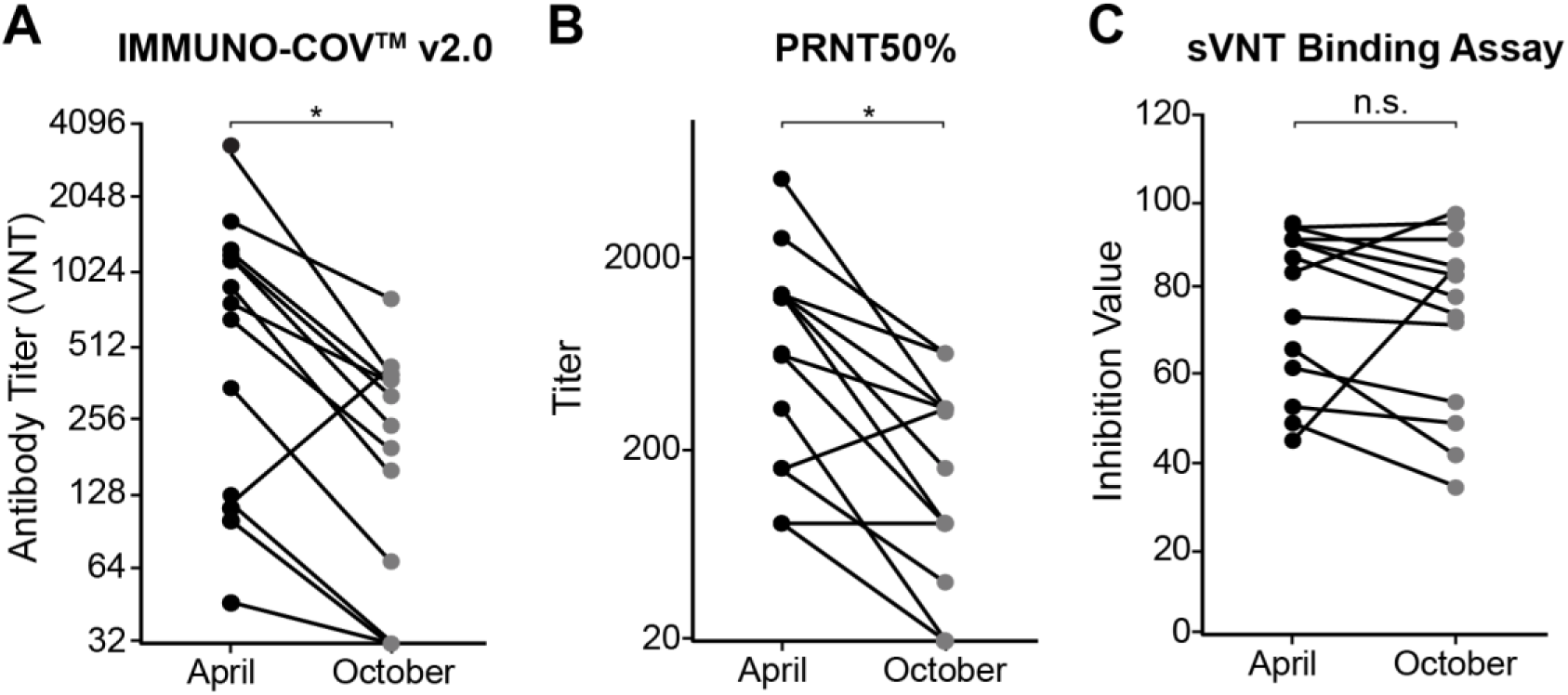
*Durability of neutralizing antibody responses*. A-C) Samples were collected from donors in April and October 2020 (n=13). Neutralizing antibody levels were measured using IMMUNO-COV™ v2.0 (A), PRNT assay (B), or the c-PASS SARS-CoV-2 neutralization antibody detection kit (C), which is a binding assay that utilizes the SARS-CoV-2 spike RBD domain. The reductions in antibody titers were statistically significant for IMMUNO-COV™ v2.0 and PRNT assay, but not for the sVNT binding assay (*p* = 0.0007, 0.0004, and 0.4669, respectively, from paired T test).

**Table 6:** Longevity of Neutralizing Antibodies

| Donor | IMMUNO-COV™ v2.0 Titer (VNT) |  |  | c-PASS Value<sup>1</sup> |  |  |
| --- | --- | --- | --- | --- | --- | --- |
|  | April<sup>2</sup> | October<sup>3</sup> | Relative titer | April<sup>2</sup> | October<sup>3</sup> | Relative value |
| 1 | 1652 | 784 | 0.47 | 94 | 95 | 1.01 |
| 2 | 1187 | 320 | 0.27 | 84 | 96 | 1.14 |
| 3 | 47 | < LOD | ≤ 0.68 | 49 | 35 | 0.71 |
| 4 | 769 | 376 | 0.49 | 92 | 92 | 1.00 |
| 5 | 3030 | 378 | 0.12 | 88 | 74 | 0.84 |
| 6 | 1179 | 246 | 0.21 | 91 | 79 | 0.87 |
| 7 | 1219 | 378 | 0.31 | 95 | 84 | 0.88 |
| 8 | 124 | < LOD<sup>4</sup> | ≤ 0.26 | 67 | 42 | 0.63 |
| 9 | 894 | 156 | 0.17 | 74 | 73 | 0.99 |
| 10 | 102 | < LOD<sup>4</sup> | ≤ 0.31 | 62 | 54 | 0.87 |
| 11 | 660 | 195 | 0.30 | 91 | 83 | 0.91 |
| 12 | 350 | 68 | 0.19 | 52 | 49 | 0.94 |
| 13 | 114 | 418 | 3.67 | 45 | 85 | 1.89 |

<sup>1</sup>Samples analyzed using the c-PASS SARS-CoV-2 Surrogate Virus Neutralization Test (sVNT) Kit
<sup>2</sup>April samples acquired from donors 2 to 8 weeks following COVID-19 symptoms or diagnosis.
<sup>3</sup>October samples acquired from same donors approximately 6 months after April samples acquired.
<sup>4</sup>Matched ACD-plasma samples were also analyzed and exhibited low-levels of neutralizing antibodies in some assay runs.

## DISCUSSION

With vaccine roll-out ongoing and critical questions still unanswered regarding the durability of protective immune responses, the need for an accurate, scalable test that can quantitatively measure SARS-CoV-2-neutralizing antibodies remains a priority. Only a small subset of antibodies capable of binding to the spike glycoprotein have neutralizing activity and are most likely to afford protection against SARS-CoV-2 infection (16,27,29,33). Commercially available monoclonal antibodies proven to be of benefit for the treatment of COVID-19 were selected based on their potent virus neutralizing activity (34–38). Yet, most serological tests currently in use detect total spike-binding antibodies but do not measure the capacity of these antibodies to neutralize virus infectivity. The traditional assay for detection and quantification of neutralizing antibodies, the PRNT, is low-throughput and for SARS-CoV-2 must be performed under high biocontainment (BSL-3), making it impractical for widespread use. Here, we describe the development and clinical validation of a novel assay, IMMUNO-COV™ v2.0, which is now available as a scalable laboratory developed test for quantitatively measuring SARS-CoV-2- neutralizing antibody titers. Our data show that IMMUNO-COV™ v2.0 can be used for accurate tracking of neutralizing antibody titers over time in individuals following natural infection or vaccination (Fig. 10). Such information will be needed to better define what constitutes a protective immune response, and what is the durability of the protective immune response following natural infection or vaccination. Answers to these questions will be important to better inform vaccine dosing schedules and other public health initiatives aimed at controlling the pandemic.

The IMMUNO-COV™ v2.0 assay measures the concentration of antibodies in serum or plasma that can neutralize the infectivity of the VSV-SARS2-Fluc virus in Vero-ACE2 cells, as detected by a reduction in luciferase activity compared to cells that have been infected in the absence of neutralizing antibodies (Fig. 1). Importantly, results from IMMUNO-COV™ v2.0 correlate closely with PRNT50% titers determined using a clinical isolate of SARS-CoV-2 (Fig. 8), indicating that neutralization of VSV-SARS2-Fluc accurately mirrors SARS-CoV-2 neutralization. Other groups have likewise observed strong correlation between the readouts of virus neutralization assays using VSV and lentiviral pseudotypes displaying the SARS-CoV-2 spike glycoprotein and readouts of classical PRNT conducted under BSL-3 using clinical isolates of SARS-CoV-2 (18,22,39). Given the strong correlations between titers determined using IMMUNO-COV™ v2.0 and those determined using classical PRNT50% and PRNT80% assays, we generated a conversion table that facilitates the rapid conversion of IMMUNO-COV™ v2.0 titers to corresponding PRNT50% titers (Table 5). Moreover, the VNT scale for IMMUNO-COV™ v2.0 was designed to yield numerical values roughly equivalent to the PRNT50% titers obtained for a given sample.

The currently available spectrum of tests for determining titers of SARS-CoV-2-neutralizing antibodies are based on clinical isolates of SARS-CoV-2 (PRNT) (40–42), replicating surrogate viruses (typically VSV-derived) (20–22), non-replicating spike protein pseudotyped viruses (primarily using VSV or lentiviruses) (10,14,17–19), or entirely nonviral platforms (RBD-ACE2 binding assays) (43, 44). Binding assays using spike receptor binding domain (RBD) are attractive due to the speed at which results can be obtained. However, they measure only that subset of neutralizing antibodies capable of blocking the binding of the SARS-CoV-2 spike protein RBD to its immobilized ACE2 receptor. They do not functionally measure virus neutralization, and since only a portion of SARS-CoV-2-neutralizing antibodies binds to the RBD (27, 29), the relevance of these assays relative to those that directly measure the inhibition of virus infection remains an open question. In relation to this important question, we observed a strong correlation between IMMUNO-COV™ v2.0 and PRNT50% titers in samples acquired at different times following SARS-CoV-2 infection. In contrast, we observed a much less robust correlation between PRNT50% titers and the c-Pass SARS-CoV-2 surrogate virus neutralization test kit, which is a spike RBD binding assay (Fig. 10).

In addition to comparing our assay to the gold standard PRNT assay, we performed full clinical validation of IMMUNO-COV2™ v2.0, which included evaluating the parameters of linearity, assay dynamic range, sensitivity, determination of the limit of blank (LOB) and limit of detection (LOD), dilutional linearity and parallelism, precision, clinical agreement, matrix equivalence, clinical specificity and sensitivity, and assay robustness. IMMUNO-COV™ v2.0 exhibited excellent clinical agreement with 100% assay specificity (Table 2). We also tested samples obtained predominately before 2019 from individuals recovered from infection with one of the four common human coronaviruses (HKU1, NL63, OC43, or 229E). All these samples tested negative for neutralizing antibodies, suggesting that IMMUNO-COV™ v2.0 is specific to SARS-CoV-2-neutralizing antibodies and most likely will not detect neutralizing antibodies directed against other human coronaviruses.

As has been reported by others (7,8,16,28,32), we observed that donors recovering from more severe COVID-19 disease generally developed higher-titer neutralizing antibody responses (Fig. 9). However, several individuals with only mild COVID-19 symptoms developed strong neutralizing antibody responses, and two individuals with severe disease developed relatively weak neutralizing antibody responses. Thus, SARS-CoV-2-neutralizing antibody titers cannot be accurately predicted based on the severity of the disease manifestations that an individual experiences, highlighting the importance of neutralizing antibody testing to determine anti-SARS-CoV-2 immune status. Irrespective of the initial magnitude of the neutralizing antibody response, repeat IMMUNO-COV™ v2.0 testing demonstrated a relatively steep decline in SARS-CoV-2-neutralizing antibody titers over six months (Fig. 10). This finding is in keeping with those of other investigators (7–11), and highlights the importance of tracking neutralizing antibodies over time. It should be noted that some other studies suggest that SARS-CoV-2- neutralizing antibody titers are relatively stable (45, 46). More research is needed to better understand the durability of neutralizing antibody responses to SARS-CoV-2 and their relationship to cell-mediated responses. Further investigation is also needed to determine whether vaccination provides immunity against SARS-CoV-2 viral variants, and we are conducting ongoing studies to confirm that IMMUNO-COV™ v2.0 can detect immunity against SARS-CoV-2 variants.

It is not currently known what minimum titer of SARS-CoV-2-neutralizing antibodies is necessary to assure protection against future infection. Likely there will be considerable variation between individuals because of the multiple additional factors impacting susceptibility to infection, including age, sex, race, ethnicity, and various comorbid conditions. Nevertheless, it is widely accepted that higher levels of neutralizing antibodies afford a higher degree of protection from future infection. Large, coordinated studies following SARS-CoV-2-neutralizing antibody titers in various cohorts of vaccinated and previously infected individuals will be needed to understand immune correlates of protection, the durability of the protective response, and the appropriate frequency for administration of booster doses of the approved SARS-CoV-2 vaccines. With the advent of IMMUNO-COV™ v2.0, a fully validated, high throughput laboratory developed test that accurately and robustly determines neutralizing antibody titers, we can now move forward with these much-needed population studies. We have generated and cryopreserved sufficient VSV-SARS2-Fluc virus to perform over 5 million assays, and the assay is accurate and reproducible even between different virus lots (Fig. 4). Moreover, during validation testing, the IMMUNO-COV™ v2.0 assay exhibited favorable precision compared to the PRNT, with acceptable levels of intra- and inter-assay variability (Table 3) and low run-to-run variability in quantitative VNT readouts. Therefore, we believe that IMMUNO-COV™ v2.0 will provide a useful and lasting standardized assay that can be used to normalize and harmonize neutralizing antibody titers for consistent monitoring of neutralizing antibody levels over time and in large study populations.

## MATERIALS AND METHODS

### Cells

African green monkey Vero cells (ATCC® CCL-81™), Vero-αHis (47), and baby hamster kidney BHK-21 cells (ATCC® CCL-10™) were maintained in high-glucose DMEM supplemented with 5% fetal bovine serum and 1X penicillin/streptomycin (complete media) at 37°C/5% CO_2_. Vero-ACE2-Puro (Vero-ACE2) cells were generated by transducing Vero cells with lentiviral vector LV-SFFV-ACE2-Puro, encoding the human ACE2 cDNA (GenBank BC039902) under control of the spleen focus forming virus (SFFV) promoter and linked to the puromycin resistance gene via a P2A cleavage peptide. Vero-ACE2-Puro/TMPRSS2-Puro (Vero-ACE2/TMPRSS2) cells were generated by transducing Vero-ACE2-Puro cells with lentiviral vector SFFV-TMPRSS2-Puro encoding human TMPRSS2 cDNA (GenBank: BC051839) under control of the SFFV promoter and linked to the puromycin resistance gene via a P2A cleavage peptide. Vectors used for stable-cell generation were verified by whole plasmid sequencing performed by MGH CCIB DNA Core (Cambridge, MA). Transduced cells were selected using 10 µg/mL puromycin. Following selection, Vero-ACE2 cells were maintained in complete media supplemented with 5 µg/mL puromycin. Puromycin was excluded when cells were seeded for assays.

### Generation of VSV-SARS2-Fluc

Full-length Luc2 (Fluc) was PCR-amplified from pLV-SFFV-Luc2-P2A-Puro (Imanis #DNA1034) with a 5’ NheI and 3’ AscI restriction site. To generate the viral genome, the amplified PCR product was cloned into pVSV-SARS-CoV-2-S-Δ19CT (21) between the SΔ19CT and L genes (Figure 2A) using the NheI and AscI restriction sites. Plasmid was sequence verified and used for infectious virus rescue on BHK-21 cells as previously described (48). VSV-G was co-transfected into the BHK-21 cells to facilitate rescue but was not present in subsequent passages of the virus. For initial amplification, the virus was propagated in Vero-αHis cells by inoculating 80% confluent monolayers in 10-cm plates with 1 mL of virus. Virus was harvested 48 h after inoculation, aliquoted, and stored at ≤ -65°C until use. For further amplifications and generation of large-scale stocks, the virus was propagated in Vero-ACE2 cells by inoculating 90% confluent monolayers at an MOI of 0.02 or 0.03 plaque forming units per cell. Virus was harvested after 48 h, aliquoted, and stored at ≤ -65°C until use. Aliquots were used to determine viral titers by plaque assay on Vero-αHis cells.

### Replication Curves

Vero-ACE2 or BHK-21 monolayers in 10-cm plates were inoculated in duplicate with OptiMEM alone (mock), VSV-Fluc (MOI=0.01), or VSV-SARS2-Fluc (MOI=0.01). After 2 h at 37°C/5% CO_2_, complete media was added to a total volume of 6 mL/ plate. At 2, 12, 24, 36, and 48 h, 0.25 mL aliquots of culture supernatant were removed from plates and replaced with 0.25 mL of fresh media. Aliquots were stored at ≤ -65°C immediately after collection until the time of titering. To determine viral titers, aliquots were thawed and assayed by plaque assay on Vero-αHis cells. Throughout the infection time course, cell photos were taken from the 10-cm plates at a 100x magnification using an inverted microscope.

### Reagents

D-luciferin potassium salt (Gold Biotechnology #LUCK-1G) was diluted in DPBS to generate 15 mg/mL stocks. For initial studies, 20 µL/well of stocks were used for assays. For later studies (starting with validation studies), stocks were diluted 1:10 in DPBS and 50 µL/well were used for assays. mAb10914 and mAb10922 are human α-SARS-CoV-2 spike neutralizing monoclonal antibodies. mAb10914 was prepared and scaled up using methods previously described by Regeneron Pharmaceuticals, Inc. (35), and mAb10922 was purchased from GenScript (#U314YFG090_1).

### Luciferase Assay Time Course

Vero-ACE2 or BHK-21 cell monolayers in 96-well black-walled plates with clear bottoms were infected with VSV-Fluc or VSV-SARS2-Fluc at a multiplicity of infection of 0.03 plaque forming units per cell. Media only wells were used as mock controls. For each condition, 24 wells were prepared to facilitate 8 time points done in triplicate. At 2, 12, 16, 20, 24, 28, 36, and 48 h after inoculation, d-luciferin was added to one set of triplicate wells and bioluminescence was immediately measured using a Tecan Infinite II instrument (100 ms integration, 100 ms settle time per well).

### Collection of Plasma and Sera Samples

A clinical protocol to collect blood samples for assay validation was reviewed and approved by Western IRB on April 1, 2020 (study ID: VYR-COV-001). Samples were obtained with informed consent and the protocol was conducted under ICH-GCP and all applicable sections of the Code of Federal Regulations. Serum and plasma samples were collected in April 2020 from patients who had previously tested positive for SARS-CoV-2 infection by a PCR test, patients who had known exposure to individuals infected with SARS-CoV-2 and symptoms of COVID-19, and a cohort of patients with no known exposure to or symptoms of COVID-19 and presumed to be seronegative. Clinical information was self-reported. A total of 150 adult volunteers were enrolled and provided blood samples at BioTrial in Newark, New Jersey and Olmsted Medical Center in Rochester, Minnesota in April 2020. A subset of 26 participants returned and volunteered a second blood sample 6 months later in October 2020.

Geisinger provided 140 frozen sera samples comprising the endemic human coronavirus panel. These samples were collected from subjects who had tested positive for the presence of coronavirus HKU1, coronavirus NL63, coronavirus OC43, coronavirus 229E using the Geisinger Respiratory Pathogen Panel PCR test (Geisinger Medical Labs) on average 282.5 days before the collection date (median: 129.3 days; range 1171.3 - 29.1 days).

### IMMUNO-COV™ v2.0 Neutralization Assays

Except where noted during initial optimization experiments, Vero-ACE2 cells were seeded at 1×10^4^ cells/well in 96-well black-walled plates with clear bottoms 16-24 h before being used for neutralization assays. On the day of assay, test samples and controls were prepared and mixed with VSV-SARS2-Fluc in U-bottom suspension cell culture plates to a final volume of 240 µL/well. Any indicated antibody concentrations or sample dilutions represent the antibody concentration or sample dilution following mix with virus. Except when noted otherwise, serum samples were thawed and used for assay without additional processing, while plasma samples were prepared by heat-inactivation for 30 min at 56°C, followed by clarification at 12,000 × g for 5 min and transfer of the liquid supernatant to fresh tubes. During initial optimization experiments, various concentrations of virus were tested, but for all subsequent assays, virus was used at 300 pfu/well (300 pfu/100 µL in U-well mixtures). Virus, test samples, and controls were all diluted as appropriate in OptiMEM to generate final concentrations. For each plate, a standard curve consisting of 0.8, 0.4, 0.2, 0.1, 0.05, and 0.025 µg/mL mAb10914 in OptiMEM, and controls NC (pooled negative matrix at 1:80), QC High (0.154 µg/mL mAb1022 in pooled negative matrix at 1:80), and QC Low (0.031 µg/mL mAb10922 in pooled negative matrix at 1:80) were included. Virus mixes in U-well plates were incubated at room temperature for 30-45 min, and then 100 µL of mixes were overlaid onto the Vero-ACE2 monolayers in duplicate. Plates were returned to a 37°C/5% CO_2_ incubator for 24-28 hours. D-luciferin was then manually added to wells using a multi-channel pipet, and luminescence was read immediately (30-90 seconds) after d-luciferin addition using a Tecan M Plex or Tecan Lume instrument (100 ms integration, 100 ms settle time per well).

### Determination of Virus Titers

Virus neutralizing titers (VNTs) were determined based on a calibration curve. The calibration curve was run on each plate and consisted of mAb10914 spiked into pooled SARS-CoV-2 seronegative sera at 0.8, 0.4, 0.2, 0.1, 0.05, and 0.025 µg/mL. From the calibration curve, the equivalent concentration of neutralizing antibody for a given luciferase signal was determined. To convert to VNT, the antibody equivalent concentration was multiplied by 400, a correction factor chosen to yield VNT values similar to PRNT50% values.

### Determinant of Limit of Blank (LOB) and Limit of Detection (LOD)

Seven known seronegative samples were analyzed at 1:80 dilution on 12 different assay runs, performed on three consecutive days, by six different analysts, using two separate virus lots. Luciferase signal relative to a media control was determined for each sample. The datasets were non-normal by Anderson-Darling and Shapiro-Wilk test, so the LOB was established using a non-parametric model with the 5^th^ percentile value of relative luciferase response obtained for each dataset. From this analysis, the LOB was a response level of 124.5%. To determine the LOD, five seronegative samples (at 1:80 dilution) were spiked with low levels of calibrator material (mAb10914) at 0.01, 0.02, 0.04, 0.06, 0.08, or 0.1 μg/mL and assayed on 12 different assay runs, performed on three consecutive days, by six different analysts, using two separate virus lots. Every run also included unspiked negative samples and media control. Datasets were evaluated for the titer that resulted in a response level below the corresponding LOB for each of the dilutions. From these analyses, the LOD was determined to be 32 VNT.

### Blinded Sample Testing

Sera and plasma samples were randomized by independent operators prior to being given to analysts for testing. Samples were assayed in batches, with an unknown number of positive and negative samples in each batch. All samples were assayed at 1:80, 1:160, 1:320, 1:640, 1:1280, and 1:2560 dilutions. For specificity and sensitivity studies, each blinded sample was tested by four different analysts, on at least three different days, in a total of five separate assay runs, using two different virus lots. For comparison studies, samples were tested using the EUROIMMUN anti-SARS-CoV-2 ELISA (IgG) according to the manufacturer’s directions.

### Assay Variability Assessment

QC High (0.154 µg/mL), QC Low (0.031 µg/mL), and matrix blank (0 µg/mL) controls consisting of mAb10922 diluted in pooled negative serum (at 1:80) were used along with the standard curve to assess assay variability. For inter-assay variability studies, controls were tested in duplicate on a total of 207 assay runs performed by five different analysts across a span of five days using two different lots of virus. For intra-assay variability studies, each control was assayed in 24 wells in the same assay run performed by the same analyst.

### Matrix Equivalency Assessment

Matched serum, sodium heparin plasma, ACD plasma, and K2/EDTA plasma samples were obtained (see Collection of plasma and sera samples). Samples were blinded and assayed as described for blinded sample testing, using appropriate pooled negative matrix controls.

### PRNT

Serum samples were heat-inactivated for 30 min at 56°C and serially 2-fold diluted in Dulbecco’s minimal essential medium supplemented with 2% heat-inactivated fetal bovine serum. SARS-CoV-2 (USA-WA1/2020) (49) was diluted to approximately 200 PFU/mL and mixed with an equal volume of diluted serum (final dilutions of serum with virus were 1:20, 1:40, 1:80, 1:160, 1:320, 1:640, 1:1280, 1:2560, 1:5120, 1:10240, 1:20480, 1:40960). Virus mixed with an equal volume of medium alone was used as a control. After a 1 h incubation at 37°C, 250 µL of virus/serum or virus/media mixes were used to inoculate Vero-E6 monolayers in 6-well plates. Absorption proceeded for 1 h at 37°C with occasional rocking, before monolayers were overlaid with 4 mL of 1.6% low-melting agarose in Minimal Essential Media supplemented with 4% fetal bovine serum and antibiotics. Plates were incubated at 37°C for two days when plaques appeared, then fixed with 10% formaldehyde, and stained with 2 mL of 0.05% neutral red, followed by incubation for 6 h at 37°C. Plaques were counted and the PRNT50% and PRNT80% titers were determined as the lowest dilution at which the number of plaques was reduced by 50% or 80%, respectively, compared to the virus/medium control. Plaque counts greater than 30 were too numerous to count and were considered as equivalent to the virus/media control.

### sVNT Binding Assay

Serum samples were tested using the SARS-CoV-2 Surrogate Virus Neutralization Test (sVNT) Kit (GenScript #L00847) according to the manufacturer’s directions.

### Flow Cytometry

Vero-αHis (Vero) or Vero-ACE2 cells were dislodged using Versene, counted, and transferred to microcentrifuge tubes (5×10^5^ cells/tube was used for ACE2 staining and 1.5×10^6^ cells/tube was used for TMPRSS2 staining). For ACE2 staining, cells were pelleted and resuspended in 100 µL FACS buffer (2% FBS in DPBS) containing 0.2 µg goat-α-human ACE2 (R&D Systems #AF933). After 30 min on ice, cells were rinsed with 1 mL FACS buffer and resuspended in 100 µL FACS buffer containing 5 µL donkey-α-goat IgG-PE secondary antibody. After 30 min on ice, cells were rinsed with 1 mL FACS buffer and fixed with 1% paraformaldehyde for 15 min on ice. Cells were washed twice with FACS buffer, resuspended in 500 µL FACS buffer and analyzed on a CYTOFLEX flow cytometer (Beckman Coulter). For TMPRSS2 staining cells were resuspended in 1 mL ice-cold 70% ethanol in DPBS and incubated on ice for 10 min. Cells were centrifuged, washed once with 1 mL FACS buffer, and resuspended in 100 µL of a 0.5% saponin solution containing 4 µg rabbit α-TMPRSS2 (Invitrogen #PA5-14264). After 30 min on ice, samples were washed twice with 1 mL FACS buffer and resuspended in 100 µL of a 0.5% saponin solution containing 2 µL goat α-rabbit IgG-AF647 secondary antibody. After 30 min on ice, cells were washed twice with FACS buffer and fixed with 1% paraformaldehyde for 15 min on ice. Cells were washed twice with FACS buffer, resuspended in 500 µL FACS buffer and analyzed on a CYTOFLEX flow cytometer (Beckman Coulter). For both ACE2 and TMPRSS2 staining, positive staining was compared against a control sample stained with secondary antibody only.

### Immunoblot

Viruses were concentrated by high-speed centrifugation, and 5 × 10^5^ pfu (VSV-SARS2-Fluc) or 5 × 10^5^ TCID50 units (VSV-GFP) were diluted in LDS sample buffer (Invitrogen #B0007) and reducing agent (Invitrogen #B0009) according to the manufacturer’s directions. Cell lysates from HEK-293T cells stably expressing SARS-CoV-2 spike protein were also prepared as controls. All samples were incubated at 70°C for 10 min and 40 µL of each sample was run in duplicate on 4-12% Bis-Tris gels (Invitrogen #NW04125Box) along with precision plus protein dual color standard (Bio-Rad #161-0374). Proteins were transferred to nitrocellulose membranes using a Power Blotter XL. Membranes were blocked in 5% non-fat dry milk in TBST, washed three times with TBST, and incubated for 1 h at room temperature with primary antibody mouse α-SARS-CoV-2 Spike (1:1000, GeneTex #GTX632604) or mouse monoclonal α-VSV-G clone 8G5F11 (1:10,000, Absolute Antibody #Ab01401-2.3). Membranes were washed three times with TBST and incubated for 1 h at room temperature with secondary antibody goat α-mouse IgG-HRP (Prometheus #20-304) at 1:20,000. Membranes were washed three times with TBST, and protein bands were developed for 2 min at room temperature using ProSignal® Dura ECL Reagent (Prometheus #20-301). Protein bands were imaged using a BioRad ChemiDoc Imaging System.

### Statistical Analyses

Descriptive statistics, comparisons, and regression analyses were performed in Graph Pad Prism, v9.0.0 (San Diego, CA). Tests for normality of variance were conducted, and whenever possible parametric comparisons were used. For non-normal datasets, non-parametric approaches were used. A four-parameter non-linear regression was used for the calibration curve of the virus neutralizing units within the assay. For correlation analyses, Spearman’s correlation analysis was conducted.

## Data Availability

A final validation summary including all the data generated as part of assay validation is in preparation. It will be disclosed to regulatory authorities and posted on public registries, as required.

## ACKNOWLEDGMENTS

We would like to thank all the participants in our clinical trial, without whose willingness to donate blood we would not have been able to develop and validate this assay. We thank DiscovEHR Study with Geisinger Health System for the generous supply of serum samples from patients recovered from endemic coronavirus infection. We would also like to thank the entire Imanis Life Sciences, Vyriad, Regeneron, BioTrial, and OMC teams, and John Mills and Elitza Thiel at Mayo Clinic for helpful discussions, laboratory, and operational support. Natalie Thornburg provided the SARS-CoV-2 isolates and Kenneth and Jessica Plante provided SARS-CoV-2 stocks, cells, and protocols. Funding for this project was provided by Regeneron Pharmaceuticals, Inc. as part of an ongoing collaboration with Vyriad, Inc., and by NIH grant AI120942 to SCW.

## AUTHOR CONTRIBUTIONS

Designed and planned experiments: RV, TC, SR, CL

Performed experiments: RV, SR, CL, RN, L. Schnebeck, AR, KS, SW, GR Analyzed data: RV, TC, SR, CL

Cloned and rescued virus: PL, CG, MH, SR

Generated critical reagents (cells, mAbs): JB, SR, A. Baum, CAK

Clinical trial implementation and test sample acquisition: A. Bexon, SN, BB, L. Suksanpaisan

Wrote the manuscript: RV, SJR

Contributed intellectually to assay development: RV, TC, SJR, L. Suksanpaisan, KWP, ST

## REFERENCES

1. Baden LR, El Sahly HM, Essink B, Kotloff K, Frey S, Novak R, et al. 2021. Efficacy and Safety of the mRNA-1273 SARS-CoV-2 Vaccine. N Engl J Med. 384:403–16.

2. Polack FP, Thomas SJ, Kitchin N, Absalon J, Gurtman A, Lockhart S, et al. 2020. Safety and Efficacy of the BNT162b2 mRNA Covid-19 Vaccine. N Engl J Med. 383:2603–15.

3. Koff WC, Burton DR, Johnson PR, Walker BD, King CR, Nabel GJ, et al. 2013. Accelerating Next Generation Vaccine Development for Global Disease Prevention. Science. 340:1232910.

4. Gao Q, Bao L, Mao H, Wang L, Xu K, Yang M, et al. 2020. Development of an Inactivated Vaccine Candidate, BBIBP-CorV, with Potent Protection against SARS-CoV-2. Cell. 182:713–721.

5. Yu J, Tostanoski LH, Peter L, Mercado NB, McMahan K, Mahrokhian SH, et al. 2020. DNA vaccine protection against SARS-CoV-2 in rhesus macaques. Science. 369:806–11.

6. Plotkin SA. 2010. Correlates of protection induced by vaccination. Clin Vaccine Immunol. 17:1055–65.

7. Lau EHY, Tsang OTY, Hui DSC, Kwan MYW, Chan W hung, Chiu SS, et al. 2021. Neutralizing antibody titres in SARS-CoV-2 infections. Nat Commun. 12:1–7.

8. Seow J, Graham C, Merrick B, Acors S, Pickering S, Steel KJA, et al. 2020. Longitudinal observation and decline of neutralizing antibody responses in the three months following SARS-CoV-2 infection in humans. Nat Microbiol. 5:1598–607.

9. Crawford KHD, Dingens AS, Eguia R, Wolf CR, Wilcox N, Logue JK, et al. 2021. Dynamics of Neutralizing Antibody Titers in the Months After Severe Acute Respiratory Syndrome Coronavirus 2 Infection. J Infect Dis. 223:197–205.

10. Brochot E, Demey B, Touzé A, Belouzard S, Dubuisson J, Schmit JL, et al. 2020. Anti-spike, Anti-nucleocapsid and Neutralizing Antibodies in SARS-CoV-2 Inpatients and Asymptomatic Individuals. Front Microbiol. 11:1–8.

11. Widge A, Rouphael NG, Jackson LA, Anderson EJ, Roberts PC, Makhene M, et al. 2021. Durability of Responses after SARS-CoV-2 mRNA-1273 Vaccination. N Engl J Med. 384:80–2.

12. Payne S. 2017. Immunity and Resistance to Viruses. Viruses. p. 61–71.

13. Nayak K, Gottimukkala K, Kumar S, Reddy ES, Edara VV, Kauffman R, et al. 2020. Characterization of neutralizing versus binding antibodies and memory B cells in COVID-19. bioRxiv. https://doi.org/10.1101/2020.08.31.276675.

14. Wu F, Wang A, Liu M, Wang Q, Chen J, Xia S, et al. 2020. Patient Cohort and Their Implications. medRxiv. https://doi.org/10.1101/2020.03.30.20047365.

15. Wu F, Liu M, Wang A, Lu L, Wang Q, Gu C, et al. 2020. Evaluating the Association of Clinical Characteristics with Neutralizing Antibody Levels in Patients Who Have Recovered from Mild COVID-19 in Shanghai, China. JAMA Intern Med. 180:1356–62.

16. Robbiani DF, Gaebler C, Frauke M, Lorenzi JC, Wang Z, Cho A, et al. 2020. Convergent Antibody Responses to SARS-CoV-2 in Convalescent Individuals. Nature. 584:437–42.

17. Crawford KHD, Eguia R, Dingens AS, Loes AN, Malone KD, Wolf CR, et al. 2020. Protocol and reagents for pseudotyping lentiviral particles with SARS-CoV-2 Spike protein for neutralization assays. bioRxiv. 10.1101/2020.04.20.051219.

18. Neerukonda SN, Vassell R, Herrup R, Liu S, Wang T, Takeda K, et al. 2020. Establishment of a well-characterized SARS-CoV-2 lentiviral pseudovirus neutralization assay using 293T cells with stable expression of ACE2 and TMPRSS2. bioRxiv. https://doi.org/10.1101/2020.12.26.424442.

19. Nie J, Li Q, Wu J, Zhao C, Hao H, Liu H, et al. 2020. Establishment and validation of a pseudovirus neutralization assay for SARS-CoV-2. Emerg Microbes Infect. 9:680–6.

20. Case JB, Rothlauf PW, Chen RE, Liu Z, Zhao H, Kim AS, et al. 2020. Neutralizing antibody and soluble ACE2 inhibition of a replication-competent VSV-SARS-CoV-2 and a clinical isolate of SARS-CoV-2. bioRxiv. https://doi.org/10.1101/2020.05.18.102038.

21. Vandergaast R, Carey T, Reiter S, Lech P, Gnanadurai C, Tesfay M, et al. 2020. Development and validation of IMMUNO-COV™: a high-throughput clinical assay for detecting antibodies that neutralize SARS-CoV-2. bioRxiv. https://doi.org/10.1101/2020.05.26.117549.

22. Dieterle ME, Haslwanter D, Bortz RH, Wirchnianski AS, Lasso G, Vergnolle O, et al. 2020. A Replication-Competent Vesicular Stomatitis Virus for Studies of SARS-CoV-2 Spike-Mediated Cell Entry and Its Inhibition. Cell Host Microbe. 28:486–96.

23. Hoffmann M, Kleine-weber H, Schroeder S, Kruger N, Herrler T, Erichsen S, et al. 2020. SARS-CoV-2 Cell Entry Depends on ACE2 and TMPRSS2 and Is Blocked by a Clinically Proven Article SARS-CoV-2 Cell Entry Depends on ACE2 and TMPRSS2 and Is Blocked by a Clinically Proven Protease Inhibitor. Cell. 181:1–10.

24. Walls AC, Park Y-J, Tortorici MA, Wall A, Mcguire AT, Veesler D. 2020. Structure, Function, and Antigenicity of the SARS-CoV-2 Spike Glycoprotein. Cell. 180:1–12.

25. Wan Y, Shang J, Graham R, Baric RS, Li F. 2020. Receptor Recognition by the Novel Coronavirus from Wuhan : an Analysis Based on Decade-Long Structural Studies of SARS Coronavirus. J Virol. 94:e00127–20.

26. Ou X, Liu Y, Lei X, Li P, Mi D, Ren L, et al. 2020. Characterization of spike glycoprotein of SARS-CoV-2 on virus entry and its immune cross-reactivity with SARS-CoV. Nat Commun. 11:1–12.

27. Brouwer PJM, Caniels TG, van der Straten K, Snitselaar JL, Aldon Y, Bangaru S, et al. 2020. Potent neutralizing antibodies from COVID-19 patients define multiple targets of vulnerability. Science. 369:643–50.

28. Legros V, Denolly S, Vogrig M, Boson B, Rigaill J, Pillet S, et al. 2020. A longitudinal study of SARS-CoV-2 infected patients shows high correlation between neutralizing antibodies and COVID-19 severity. medRxiv. https://doi.org/10.1101/2020.08.27.20182493.

29. Liu L, Wang P, Nair MS, Yu J, Rapp M, Wang Q, et al. 2020. Potent Neutralizing Antibodies Directed to Multiple Epitopes on SARS-CoV-2 Spike. BioRxiv. https://doi.org/10.1101/2020.06.17.153486.

30. Cao Y, Su B, Guo X, Sun W, Deng Y, Bao L, et al. 2020. Potent Neutralizing Antibodies against SARS-CoV-2 Identified by High-Throughput Single-Cell Sequencing of Convalescent Patients’ B Cells. Cell. 182:73–84.

31. Yuan M, Liu H, Wu NC, Wilson IA. 2020. Recognition of the SARS-CoV-2 receptor binding domain by neutralizing antibodies. Biochem Biophys Res Commun. https://doi.org/10.1016/j.bbrc.2020.10.012.

32. Liu L, To KK-W, Chan K-H, Wong Y-C, Zhou R, Kwan K-Y, et al. 2020. High neutralizing antibody titer in intensive care unit patients with COVID-19. Emerg Microbes Infect. 9:1–30.

33. Zost SJ, Gilchuk P, Chen RE, Case JB, Reidy JX, Trivette A, et al. 2020. Rapid isolation and profiling of a diverse panel of human monoclonal antibodies targeting the SARS-CoV-2 spike protein. Nat Med. 26: 1422–7.

34. Chen P, Nirula A, Heller B, Gottlieb RL, Joseph B, Morris J, et al. 2021. SARS-CoV-2 Neutralizing Antibody LY-CoV555 in Outpatients with Covid-19. N Engl J Med. 384:229–37.

35. Hansen J, Baum A, Pascal KE, Russo V, Giordano S, Wloga E, et al. 2020. Studies in humanized mice and convalescent humans yield a SARS-CoV-2 antibody cocktail. Science. 1014:1010–4.

36. Baum A, Fulton BO, Wloga E, Copin R, Pascal KE, Russo V, et al. 2020. Antibody cocktail to SARS-CoV-2 spike protein prevents rapid mutational escape seen with individual antibodies. Science. 1018:1014–8.

37. Weinreich DM, Sivapalasingam S, Norton T, Ali S, Gao H, Bhore R, et al. 2021. REGN-COV2, a Neutralizing Antibody Cocktail, in Outpatients with Covid-19. N Engl J Med. 384: 238–51.

38. Jones BE, Brown-Augsburger PL, Corbett KS, Westendorf K, Davies J, Cujec TP, et al. 2020. LY-CoV555, a rapidly isolated potent neutralizing antibody, provides protection in a non-human primate model of SARS-CoV-2 infection. bioRxiv. https://doi.org/10.1101/2020.09.30.318972.

39. Oguntuyo KY, Stevens CS, Hung CT, Ikegame S, Acklin JA, Kowdle SS, et al. 2020. Quantifying absolute neutralization titers against SARS-CoV-2 by a standardized virus neutralization assay allows for cross-cohort comparisons of COVID-19 sera. medRxiv. https://doi.org/10.1101/2020.08.13.20157222.

40. Okba NMA, Müller MA, Li W, Wang C, GeurtsvanKessel CH, Corman VM, et al. 2020. Severe Acute Respiratory Syndrome Coronavirus 2−Specific Antibody Responses in Coronavirus Disease Patients. Emerg Infect Dis J. 26:1478–88.

41. Wang C, Li W, Drabek D, Okba NMA, van Haperen R, Osterhaus ADME, et al. 2020. A human monoclonal antibody blocking SARS-CoV-2 infection. Nat Commun. 11:1–6.

42. Perera RAPM, Mok CKP, Tsang OTY, Lv H, Ko RLW, Wu NC, et al. 2020. Serological assays for severe acute respiratory syndrome coronavirus 2 (SARS-CoV-2), March 2020. Eurosurveillance. 2:1–9.

43. Abe KT, Li Z, Samson R, Samavarchi-Tehrani P, Valcourt EJ, Wood H, et al. 2020. A simple protein-based surrogate neutralization assay for SARS-CoV-2. JCI Insight. 5:e142362.

44. Tan CW, Chia WN, Qin X, Liu P, Chen MI-C, Tiu C, et al. 2020. A SARS-CoV-2 surrogate virus neutralization test based on antibody-mediated blockage of ACE2–spike protein–protein interaction. Nat Biotechnol. 38: 1073–8.

45. Dan JM, Mateus J, Kato Y, Hastie KM, Yu ED, Faliti CE, et al. 2021. Immunological memory to SARS-CoV-2 assessed for up to 8 months after infection. Science. 4063:1–23.

46. Wajnberg A, Amanat F, Firpo A, Altman DR, Bailey MJ, Mansour M, et al. 2020. Robust neutralizing antibodies to SARS-CoV-2 infection persist for months. Science. 370:1227– 30.

47. Nakamura T, Peng KW, Harvey M, Greiner S, Lorimer IAJ, James CD, et al. 2005. Rescue and propagation of fully retargeted oncolytic measles viruses. Nat Biotechnol. 23:209–14.

48. Ammayappan A, Nace R, Peng K-W, Russell SJ. 2013. Neuroattenuation of Vesicular Stomatitis Virus through Picornaviral Internal Ribosome Entry Sites. J Virol. 87:3217–28.

49. Harcourt J, Tamin A, Lu X, Kamili S, Sakthivel SK, Murray J, et al. 2020. Severe Acute Respiratory Syndrome Coronavirus 2 from Patient with 2019 Novel Coronavirus Disease, United States. Emerg Infect Dis. 26:1266–73.

